# Sequential application of time-stratified demographic, vital, clinical-laboratory and microbiology variables for accurate and rapid identification of sepsis

**DOI:** 10.64898/2026.05.27.26354135

**Authors:** Krupa A. Navalkar, José Garnacho-Montero, María Luisa Cantón-Bulnes, José Luís García-Garmendia, Ángel Estella, Adela Fernández-Galilea, Isidro Blanco, Maria Antonia Estecha-Foncea, Marina Gordillo-Resina, Jorge Rodríguez-Gómez, Juan Jesús Pineda-Capitán, Carmen Martínez-Fernández, Ana Escoresca-Ortega, Rosario Amaya-Villar, Juan Mora-Ordóñez, Sara González-Soto, Antonio Gutierrez-Pizarraya, Robert Balk, Russell R. Miller, John P. Burke, Gourang Patel, Jorge P. Parada, Marcus J. Schultz, Brendon P. Scicluna, Emily Blodget, Santhi Kumar, Dayle Sampson, Thomas D. Yager, Roy F. Davis, Silvia Cermelli, Richard B. Brandon

## Abstract

**Background:** Accurate early identification of sepsis remains a major clinical challenge due to its heterogeneous presentation and overlap of clinical signs with the non-infectious systemic inflammatory response syndrome (SIRS). Timely differentiation is crucial for improving patient outcomes, meeting sepsis bundle requirements and reducing inappropriate antimicrobial use. We hypothesized that clinical-laboratory data available within the first 3 hours of patient presentation could be used to identify patients with sepsis to an actionable level of accuracy, in lieu of traditional microbiology results which would not become available until at least 12-24 hours. Data from two independent studies were used to quantify the diagnostic value of demographic, vital, clinical-laboratory, and microbiological data available at three time points for distinguishing retrospectively diagnosed critically ill patients with either sepsis or non-infectious SIRS. A particular focus of this work was an assessment of the utility of SeptiCyte RAPID (Immunexpress Inc., Seattle, Washington, USA) as an aid to sepsis diagnosis, producing actionable data within 1 hour.

**Methods:** Data from two independent study cohorts were analysed. The “510k cohort” consisted of 419 adult patients in intensive care (ICU) (MARS, VENUS, and NEPTUNE studies). The “Andalusian cohort” consisted of 353 ICU patients from the PANGEA study. Logistic regression models, selected by a greedy search algorithm and validated by repeated cross-validation, were used to determine the contributions of different variables to diagnostic accuracy. Diagnostic performance was quantified by area under the receiver operating characteristic curve (AUC).

**Results:** For the 510k cohort, a baseline AUC of 0.69–0.73 was observed using 5-7 vital and demographic variables assessed immediately upon presentation (time T1). The addition of clinical-laboratory variables, in particular SeptiCyte RAPID, within 1-3 hours post-presentation (time T2) increased the AUC to 0.83-0.85). Finally, the addition of microbiological data 12-24 hours post-presentation (time T3) further improved the AUC to 0.90–0.91. Similar results were obtained for the Andalusian cohort. AUC values at the three time points were as follows: At time T1, AUC = 0.67 based solely on vital signs and demographics; at time T2, AUC = 0.87 based on vitals + demographics + SeptiCyte RAPID ± other clinical-laboratory data; at time T3, AUC = 0.93 based on vitals + demographics + SeptiCyte RAPID ± other clinical-laboratory data + microbiology results). For both cohorts, the most significant variables included temperature, mean arterial pressure, respiratory rate, suspected infection site; SeptiCyte RAPID, procalcitonin, confirmed bacterial infection and positive blood culture confirmation.

**Conclusions:** Accuracy of identification of sepsis increases markedly as demographics and vital signs are supplemented with clinical-laboratory information, and ultimately with microbiological culture results. The AUC improves significantly from T1 to T2 as laboratory data, and in particular SeptiCyte RAPID results, become available. Integrating rapid host-response testing with SeptiCyte RAPID into time-based diagnostic frameworks may enhance early sepsis recognition, improve antimicrobial stewardship, and support guideline-driven clinical decisions.

## 1. Introduction

Misdiagnosis, overdiagnosis, and under-diagnosis contribute substantially to healthcare costs across Organization for Economic Cooperation and Development (OECD) nations, with diagnostic errors estimated to cause serious harm in nearly 10% of U.S. cases annually (OECD, 2025; Newman-Toker et al., 2024) [1,2]. Sepsis, a dysregulated host response to infection leading to life-threatening organ dysfunction, remains a major diagnostic challenge due to its heterogeneous presentation and overlap of clinical signs with non-infectious inflammatory conditions including the systemic inflammatory response syndrome (SIRS) [3,4] (Meyer & Prescott, 2024; Lengquist et al., 2024). Clinical signs of sepsis are often nonspecific, varying with infection site, pathogen, and patient comorbidities, and up to one-third of confirmed sepsis cases lack an identifiable infectious source [3] (Meyer & Prescott, 2024).

Diagnostic uncertainty in cases of suspected sepsis may lead to delays in targeted treatment. Consequences of treatment delays include increased hospital readmission rates, long-term cognitive and physical impairments, and increased mortality [5] (Evans et al., 2021). Misdiagnosis at admission is common (∼12%), with misdiagnosed patients having more than double the odds of death compared to correctly diagnosed patients (adjusted Odds Ratio ≈ 2.7–3.0) [6] (Abe et al., 2019). These authors concluded that early accurate diagnosis, rather than solely rapid treatment, should be a critical focus in sepsis care, as misdiagnosis contributes to worse outcomes despite adherence to time-sensitive treatment bundles.

Against this backdrop, it is clear that improving the accuracy and precision of early sepsis diagnosis will help to ensure the appropriate and timely administration of bundles, thus potentially leading to better patient outcomes. We hypothesized that clinical-laboratory variables available within the first 3 hours of patient presentation could be used to identify patients with sepsis to an actionable level of accuracy, in lieu of traditional microbiology results which will not become available until at least 12-24 hours. We present a conceptual framework for the sequential accumulation of diagnostic information, and apply this framework to the analysis of two independent datasets collected from critically ill adults in ICU.

According to this conceptual framework, a diagnostic workup for sepsis is characterized by the sequential temporal availability of data. At time T1 (patient presentation), the only information available is demographic (including patient history) and vital signs. At time T2 (1-3 hours after presentation) clinical-laboratory data, including SeptiCyte RAPID test results, become available. Finally, at time T3 (typically 1-3 days post-presentation) microbiological culture results become available. By analyzing data from two independent cohorts of ICU patients, we estimated the diagnostic performance of individual variables, and combinations of variables, at times T1, T2, T3 for distinguishing sepsis from non-infectious SIRS. The overall aim of this work was to identify key differentiating variables to assist clinicians in more accurate and timely identification of sepsis.

## 2. Materials and Methods

### 2.1 Study Cohorts / Datasets

Datasets from two independent study cohorts were used, to assess the utility of the conceptual framework (sequential data utilization at times T1, T2, T3). Although the two datasets could not be combined (due to incomplete overlap of variables), the separate analyses were largely concordant and supportive of each other.

510k Cohort (Dataset #1): This cohort included 419 critically ill adult patients, suspected sepsis within the first 24 hours of ICU admission and displaying at least two SIRS criteria. Final adjudicated diagnoses were sepsis (n = 176) or SIRS (n = 243), as described below. The cohort comprised a retrospective sub-cohort (n = 356) and a prospective sub-cohort (n = 63), as previously reported by Balk et al. (2024) [7]. The retrospective sub-cohort was drawn from the observational MARS, VENUS, and VENUS Supplement trials (NCT01905033 and NCT02127502; clinicaltrials.gov (accessed on 17 February 2024)). The recruitment dates were January 2011–December 2013 (MARS, Europe), May 2014–April 2015 (VENUS, USA) and March–August 2016 (VENUS Supplement, USA). The prospective sub-cohort consisted of 63 critically ill adult subjects enrolled in an observational trial (NEPTUNE, NCT05469048; clinicaltrials.gov (accessed on 17 February 2024)) between the dates 26 May 2020 and 25 April 2021 at Emory University/Grady Memorial Hospital (Atlanta, GA, USA), Rush University Medical Center (Chicago, IL, USA) and University of Southern California (USC) Medical Center (with two separate sites, Keck Hospital of USC, and Los Angeles General Medical Center, in Los Angeles, CA, USA).

Ethics approval for the MARS trial was given by the Medical Ethics Committee of the Amsterdam Medical Center (approval # 10-056C). Ethics approvals for the VENUS trial were given by the relevant Institutional Review Boards as follows: Intermountain Medical Center/Latter Day Saints Hospital (approval # 1024931); Johns Hopkins Hospital (approval # IRB00087839); Rush University Medical Center (approval # 15111104-IRB01); Loyola University Medical Center (approval # 208291); Northwell Healthcare (approval #16-02-42-03). Ethics approvals for the NEPTUNE trial were given by the relevant Institutional Review Boards as follows: Emory University (approval # IRB00115400); Grady Memorial Hospital (approval # 00-115400); Rush University Medical Center (approval # 19101603-IRB01); University of Southern California Medical Center (approval # HS-19-0884-CR001). All subjects, or their legally authorized representatives, gave informed consent for participation in the respective study. All methods used in this study were carried out in accordance with the relevant guidelines and regulations.

Andalusian Cohort (Dataset #2): This cohort comprised 353 critically ill ICU patients with final adjudicated diagnoses of sepsis (n = 267) or SIRS (n = 86), as described below. It was derived from the multicenter prospective PANGEA study, details of which have been published (Cantón-Bulnes et al., 2026) [8]. The study was conducted between March 3 and December 20, 2022, across seven ICUs in Andalusia, Spain, and was coordinated by the Virgen Macarena University Hospital in Seville. Two of the hospitals (Hospital Virgen del Rocío (Seville), Hospital Universitario Regional de Málaga (Málaga)) initiated their studies late (in September and October 2022, respectively) due to organizational problems and delay in the approval by the Research Ethics Committees. The study was not registered in any public clinical trial database, as it was an observational diagnostic accuracy study with no therapeutic intervention. The study followed the Standards for Reporting of Diagnostic Accuracy (STARD) guidelines for reporting studies of diagnostic accuracy. All patients aged 18 years or older, admitted to the ICU and with a diagnosis of sepsis according to the Sepsis-3 definition were included (n=353). Subjects were excluded if they were pregnant, or the clinical picture suggestive of sepsis had started more than 48 hours previously. Blood samples were collected in PAXgene Blood RNA tubes, from which 0.9mL aliquots were pipetted into SeptiCyte RAPID cartridges and run on the Idylla system onsite.

The study was approved by the Research Ethics Committees (CEI) of the Virgen Macarena-Virgen Rocio University Hospitals on 20 December 2021 (Internal Code, 2662-N-21). The research ethics committees allowed blood sample extractions prior to obtaining written permission, if needed, to enable SeptiCyte RAPID results to be generated as quickly as possible. Written consent from the patient or next of kin was obtained within 48 hours of ICU admission and the blood sample was discarded if written consent was not obtained.

### 2.2 Reference Diagnoses (Comparators)

510k Cohort: The diagnostic performance of individual and combined variables was evaluated by comparison to a ‘gold standard’ of retrospective sepsis / SIRS clinical adjudication. The adjudication process, termed retrospective physician diagnosis (RPD), was conducted under the sepsis-2 and sepsis-3 definitions and has been previously described (Miller et al., 2018) [9]. RPD used a panel of three independent expert physicians who did not attend the patients. Each physician was provided with the same information, including selected clinical and laboratory results, and were asked to provide a diagnostic call of either sepsis or SIRS (forced diagnosis). The RPD panel was blinded to SeptiCyte RAPID and procalcitonin (PCT) results to prevent the panelists’ diagnoses from being influenced by these two variables. For other variables, such as some vital signs and laboratory variables, an incorporation bias effect is acknowledged and discussed in detail below. These variables were part of the standard clinical data collected in patient charts that the RPD panelists reviewed to make their diagnosis.

Andalusian Cohort: Patients were categorized as “sepsis” or “sterile inflammation” by two investigators at each participating site who were aware of the clinical, analytical, and microbiological data but blinded to the SeptiCyte results. A Steering Committee of four clinicians reviewed all cases and contacted local investigators in case of doubt. All patients were followed up until death or hospital discharge. As for the 510k cohort, an incorporation bias effect is acknowledged for some of the variables, and discussed in detail below.

### 2.3 Variables Used in the Analyses

Variables were grouped into three categories based on their typical availability after patient presentation. Time T1, available immediately upon presentation: patient history and vital signs. Time T2, available within 1–3 hours: clinical-laboratory variables. Time T3, available within 1–3 days: microbiological culture results. Viral PCR/antigen tests (to detect active infections) were run for some but not all patients, based on individual assessment by the care team. The timing of when these tests were ordered and run was not recorded, and likely might fall beyond the 1–3-hour T2 window. Because of this timing uncertainty, viral PCR/antigen test results, when available, were included in the later T3 time point.

#### 510k Cohort

From an initial assessment of 47 different clinical-laboratory variables, 42 of the variables had less than 33.4% missing values and were included in further analyses (see Supplement Table S1). A missing value cutoff of ∼33% was based on a balance between the ability to appropriately manage missing values using imputation methods [10] (Jakobsen et al., 2017) and incorporating as many variables as possible. Missing values for 27 of these 42 variables were imputed as the means of the available values, using the function imputePCA from the missMDA R package by Josse and Husson (2016) [11].

Maximum and minimum values for the clinical laboratory variables MAP, temperature, HR, WBC, and glucose were recorded over the first 24 hours in ICU. These max and min values were used as proxies for data that would ordinarily, in clinical practice, be collected within the first few minutes of presentation (at time T1). Certain variables were binarized (e.g., respiratory rate below or above 22/min), while others were converted to numerical scales (e.g., SIRS criteria, ranging from 0 to 4). Both the total SOFA score and its individual component scores were included. Microbiological data captured the presence of bacterial, viral, or fungal infections as documented in clinical notes.

#### Andalusian Cohort

From an initial assessment of 38 different clinical-laboratory variables, three had less than 33% missing values and were included in further analyses (see Supplement Table S2). Using these 38 variables, we performed Greedy searches to identify key variables and to estimate performance.

For both cohorts, the suspected ‘site of infection’ refers to the attending clinician’s initial assessment and was considered to fall into one of seven mutually exclusive categories: pulmonary, abdominal, urinary, bloodstream, central nervous system, other, or ‘unknown’ (unknown or not recorded). For each patient, the possibilities for site of infection were coded as either “1” (suspected), or “0” (not suspected). See Supplement Table S1 for additional detail.

### 2.4 SeptiCyte RAPID

SeptiCyte RAPID (Immunexpress Inc., Seattle, Washington, USA) is a transcription-based host response test for discriminating sepsis vs. SIRS. It has received regulatory clearance for clinical use in the USA, Europe and Australia. The test is based on quantitation of transcripts from the PLAC8 and PLA2G7 genes. The PLAC8 gene (placental associated 8) encodes a protein with diverse roles in cell proliferation, immune function, and development, including its original identification in placenta-specific tissue [12] (Mao et al., 2021). PLAC8 is up-regulated in sepsis [13] (Zhang et al. 2024). The PLA2G7 (phospholipase A2 Group VII) gene encodes a protein that degrades platelet-activating factor, which is a pro-inflammatory lipid [14] (Candels et al., 2022). PLA2G7 is down-regulated in sepsis [15] (Trimoreau et al., 2000).

SeptiCyte RAPID is run on the Biocartis Idylla hardware platform, using version 1.2 of the test specific software (Biocartis NV, Mechelen, Belgium). The test is performed by pipetting 0.9 mL of PAXgene-stabilized blood (corresponding to 0.24 mL of drawn blood) into a custom cartridge, which performs all assay steps including sample extraction/purification and RT-qPCR for the detection and relative quantification of the PLAC8 and PLA2G7 mRNA targets. Test results are calculated and presented automatically through a software-generated report, which includes a quantitative score (SeptiScore, range 0–15, higher scores correlate to increased likelihood of sepsis), calculated as the difference between the RT-qPCR Cq values for PLA2G7 and PLAC8 and proportional to sepsis probability. The test has a hands-on time of ∼2 min and a turnaround time of ∼1 h.

### 2.5 Statistical Methods

#### General Statistical Analyses

As a measure of test performance, we used area under the receiver operating characteristic curve (AUC). In accordance with general practice, we consider AUC 0.80 to be the lower threshold for “good” diagnostic performance [16], as indicated by the red horizontal lines at 0.80 in Figures 2-4 below. Calculations were conducted with the pROC package (version 1.19.0.1) using the methods roc or auc, implemented in R by Robin X. et al. (2011) [17].

#### Greedy search

A strategy that combines variables in order of decreasing contribution strength is referred to as a greedy search. The algorithm was implemented as described by Huang (2003) [18]. Optimal logistic combinations of variables for discriminating sepsis from SIRS in either the 510k or Andalusian datasets were identified using this approach. Robust variable selection was achieved through internal two-fold cross-validation repeated 100 times. Model performance was evaluated using a logistic regression decision rule, with the objective of maximizing the mean area under the receiver operating characteristic curve (AUC) across cross-validation iterations.

### 2.6 Assessment of Incorporation Bias

A range of clinical and laboratory variables was considered by the RPD panelists in establishing the comparator (reference) diagnoses of sepsis or SIRS. This introduced the potential for incorporation bias, since some of these same variables were later used to assess diagnostic performance. SeptiCyte RAPID and PCT, however, were not subject to incorporation bias, because the RPD panelists were blinded to these two variables. To assess the potential influence of incorporation bias, we performed a variable-exclusion sensitivity analysis. A logistic regression model was constructed using seven informative variables identified by a greedy search (SeptiCyte RAPID, PCT, Tmax, RRmax, MAPmax, age, HRmin). AUC were calculated, and AUC confidence intervals were estimated using nonparametric bootstrap resampling (2,000 iterations) with percentile-based 95% intervals. We examined the change in AUC resulting from setting individual logistic regression coefficients, as well as selected combinations of coefficients, to zero. This analysis characterized the dependence of the sepsis/SIRS discrimination on each model component. A table and a figure in the Supplementary material present the results of these analyses.

## 3. Results

### 3.1. Conceptual Framework for the Analysis

Figure 1 presents a conceptual framework for our analysis, and a top-level summary of results from the independent 510k and Andalusian cohorts. Variables with < 33% missing values were grouped into three categories based on their typical availability after patient presentation: patient history, demographics, and vital signs, available immediately upon presentation (time T1); clinical-laboratory variables, available within 1–3 hours (time T2); and microbiology variables, available within 1–3 days (time T3). For a full list of variables for the 510k Andalusian cohorts, see Supplementary Tables S1 and S2 respectively.

**Figure 1.**
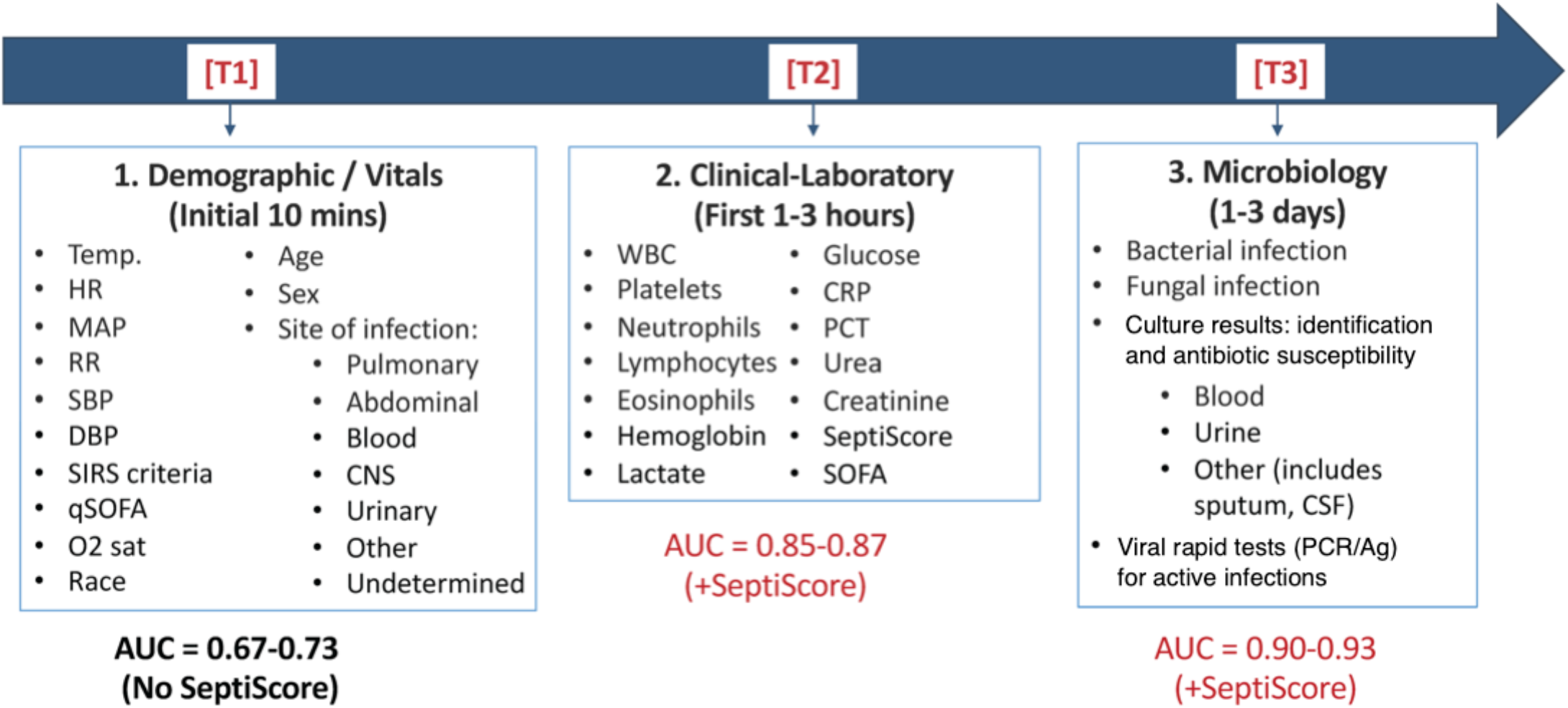
Diagnostic Variables Measured in the 510k and Andalusian cohorts. Variables can be classified according to their temporal availability. Time T1, immediately upon presentation: patient history, demographics, vital signs. Time T2, within 1–3 hours: clinical-laboratory variables including SeptiCyte RAPID. Time T3, within 1-3 days: Microbiological culture results. Viral PCR/antigen test results, when available, were assigned to the later T3 time point because the exact timing of testing was uncertain. The indicated AUC values are summarized from analyses of the 510k and Andalusian cohorts. Abbreviations: Heart Rate (HR), Respiratory Rate (RR), Mean Arterial Pressure (MAP), Systolic Blood Pressure (SBP), Diastolic Blood Pressure (DBP), Antigen (Ag), Central Nervous System (CNS), Cerebrospinal Fluid (CSF), Polymerase Chain Reaction (PCR), Procalcitonin (PCT), quick Sequential Organ Failure Assessment (qSOFA), Sequential Organ Failure Assessment (SOFA), Systemic Inflammatory Response Syndrome (SIRS).

### 3.2. Analysis Based on Data from Time T1 (Patient History, Demographics, Vital Signs)

We first considered the variables available immediately upon patient presentation which would be used in a clinical appraisal. Greedy searches were conducted to identify the most important variables for differentiating sepsis and SIRS. Figure 2 shows the results of these searches, performed (A) when a site of infection was not known, and (B) when a pulmonary site of infection was suspected. (Figures 2-4 in the main body of this paper focused on a suspected pulmonary site of infection, because this is the most common infection site in patients retrospectively determined to have sepsis [19] (Cecconi et al., 2018).) When a site of infection was not known, the maximum AUC was obtained using five variables (AUC 0.69, with 95% CI: 0.64-0.74). When the suspected site of infection was pulmonary the maximum AUC was obtained using seven variables (AUC 0.73, with 95% CI: 0.69-0.77). Plots for other common sites of suspected infection (max AUC 0.69-0.73) are included in the Supplementary Material.

Variables that consistently contributed to AUC across suspected infection sites included Temperature (Max), site of infection (if known), respiratory rate (Max), age, heart rate (Min) and Mean Arterial Pressure (Max and Min). Temperature (Max) was the single most important variable contributing to AUC across all greedy searches. The suspected site of infection was the second most important variable (adding ∼0.04 AUC units) for pulmonary, abdominal and urinary infections, and was least important for CNS infections. Contrary to intuition, respiratory rate (Max) did not contribute to AUC when the suspected site of infection was respiratory.

**Figure 2.**
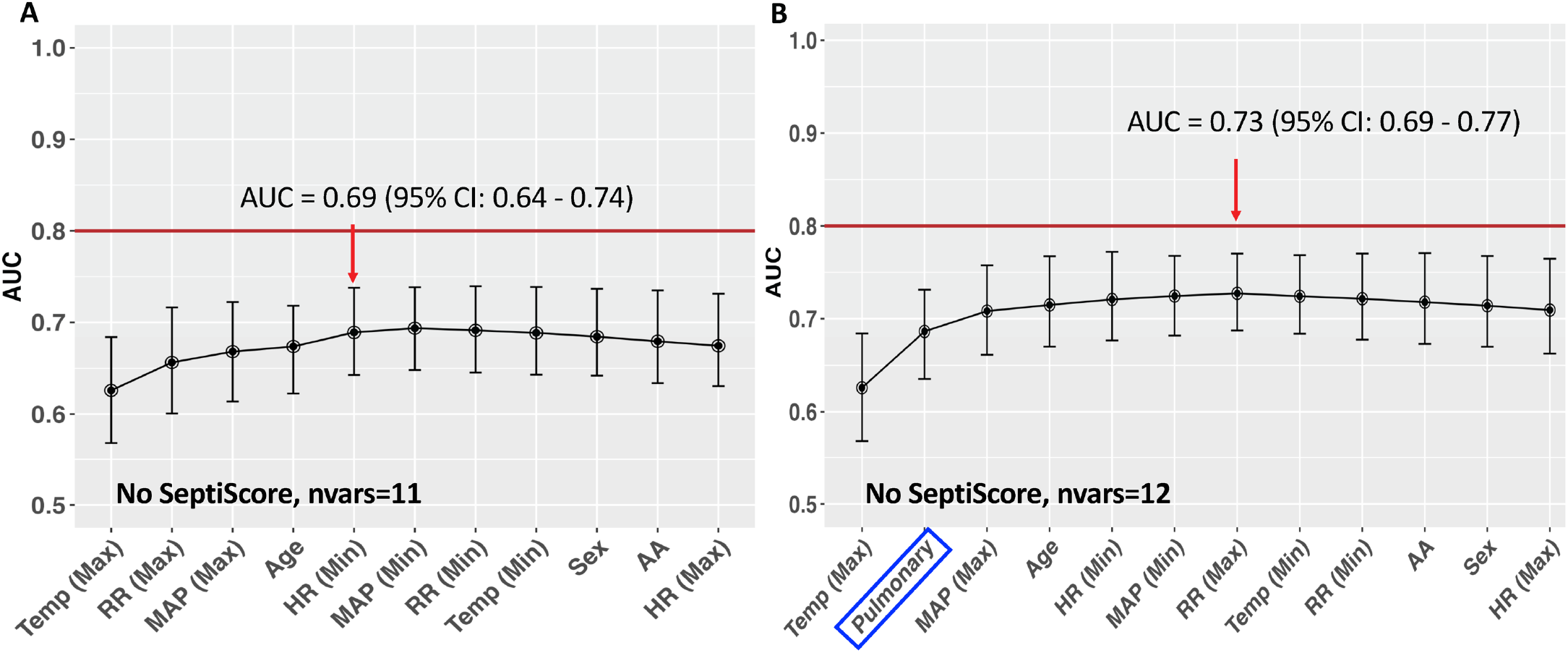
Diagnostic Performance at Time T1 (immediately upon presentation), based on patient history, demographics, vital signs. Data from the 510k cohort. (A) Site of infection not known; (B) Pulmonary site of infection suspected. Abbreviations: Temperature (Temp), Arterial (Art), Minimum (Min), Maximum (Max), AA (African American). Each dot and whisker represent the mean AUC ± 95% CI for the corresponding variable.

### 3.3. Addition of Clinical-Laboratory Data (Time T2: 1-3 hours)

For evaluation at 1–3 hours, a large number of clinical-laboratory variables become available. Figure 3 shows the greedy search results for a pulmonary site of infection, when SeptiCyte RAPID results were excluded (panel A) or included (panel B). Plots for all other suspected sites of infection are included in the Supplementary Material.

**Figure 3.**
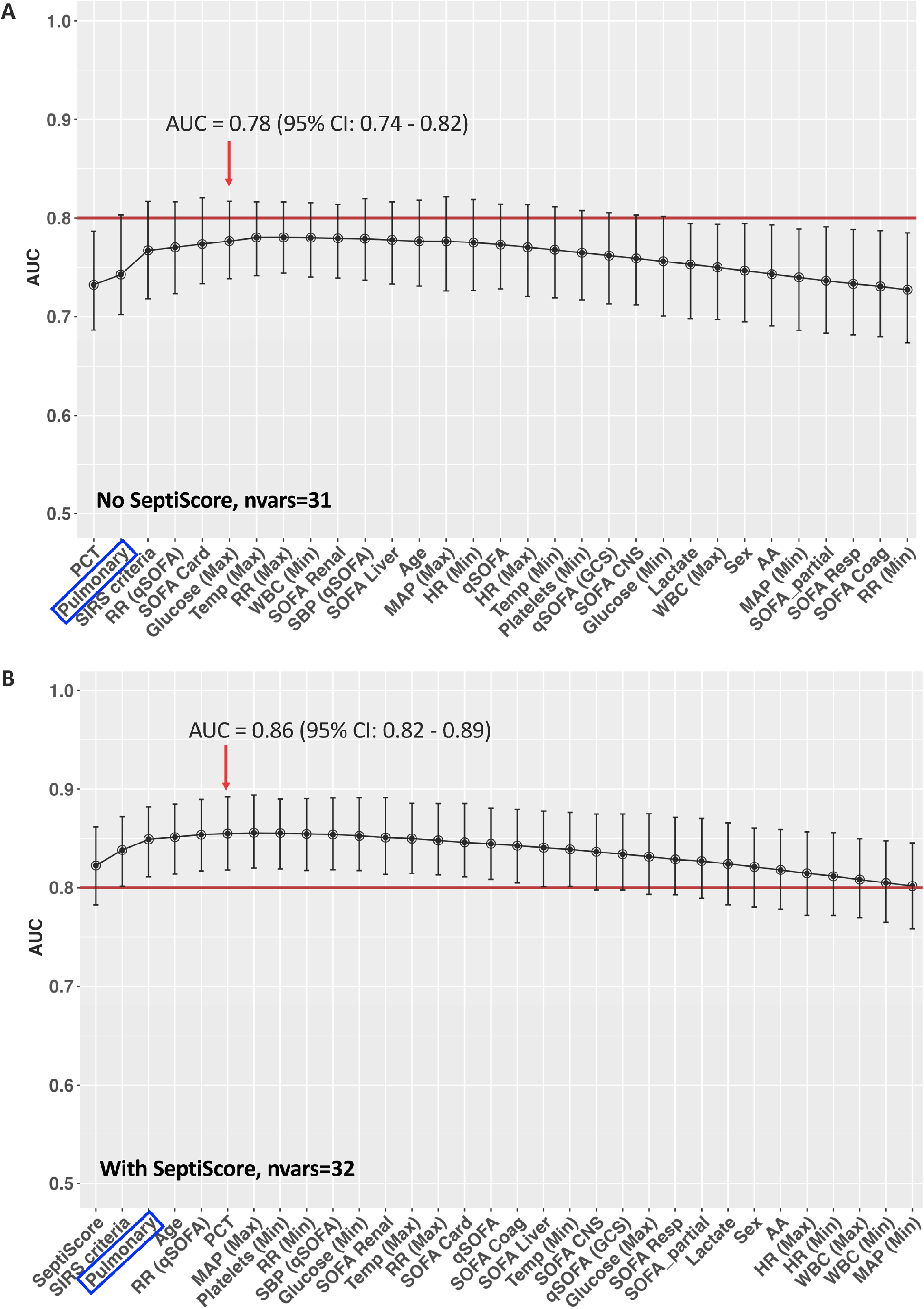
Diagnostic performance based on T1 parameters, plus clinical-laboratory test results from time T2 (1-3 hours). Analysis for Pulmonary site of infection. Data from the 510k cohort. (A) Without SeptiCyte RAPID. (B) With SeptiCyte RAPID. Abbreviations: Procalcitonin (PCT), Systemic Inflammatory Response Syndrome (SIRS), quick Sequential Organ Failure Assessment (qSOFA), Respiratory Rate (RR), Sequential Organ Failure Assessment (SOFA), Maximum (Max), Minimum (Min), Glasgow Coma Score (GCS), White Blood Cell Count (WBC), Respiratory (Resp), Coagulation (Coag), Arterial (Art). Each dot and whisker represent the mean AUC ± 95% CI for the corresponding variable.

*When excluding SeptiCyte RAPID*, the maximum AUC was 0.78 (95% CI: 0.74-0.82), which was achieved with six variables (Figure 3A). The variables that consistently contributed the most to AUC across all searches were PCT, site of infection, and SIRS criteria. Procalcitonin (PCT) was the single most important variable across all greedy searches when SeptiCyte RAPID was not included.

*When including SeptiCyte RAPID*, the maximum AUC was 0.86 (95% CI: 0.82-0.89), which was achieved with six variables (Figure 3B). the variables that consistently contributed the most to AUC across all searches were SeptiScore, site of infection, and SIRS criteria. SeptiCyte RAPID was the single most important variable that contributed to AUC across all greedy searches and was not synergistic with PCT.

### 3.4. Addition of Microbiological Culture Results (Time T3: 1-3 days)

For assessment at 1–3 days, microbiological culture results become available. Figure 4 summarizes the diagnostic performance that could be obtained by combining test results from T1 (time of presentation), T2 (clinical-laboratory data at 1-3 hours), and T3 (microbiological culture results at 1-3 days). Without SeptiCyte RAPID, the AUC could be increased to 0.87 (95% CI: 0.84-0.90) using seven variables (panel A). With SeptiCyte RAPID, the AUC could be increased to 0.90 (95% CI: 0.87-0.93) with six variables (panel B).

**Figure 4.**
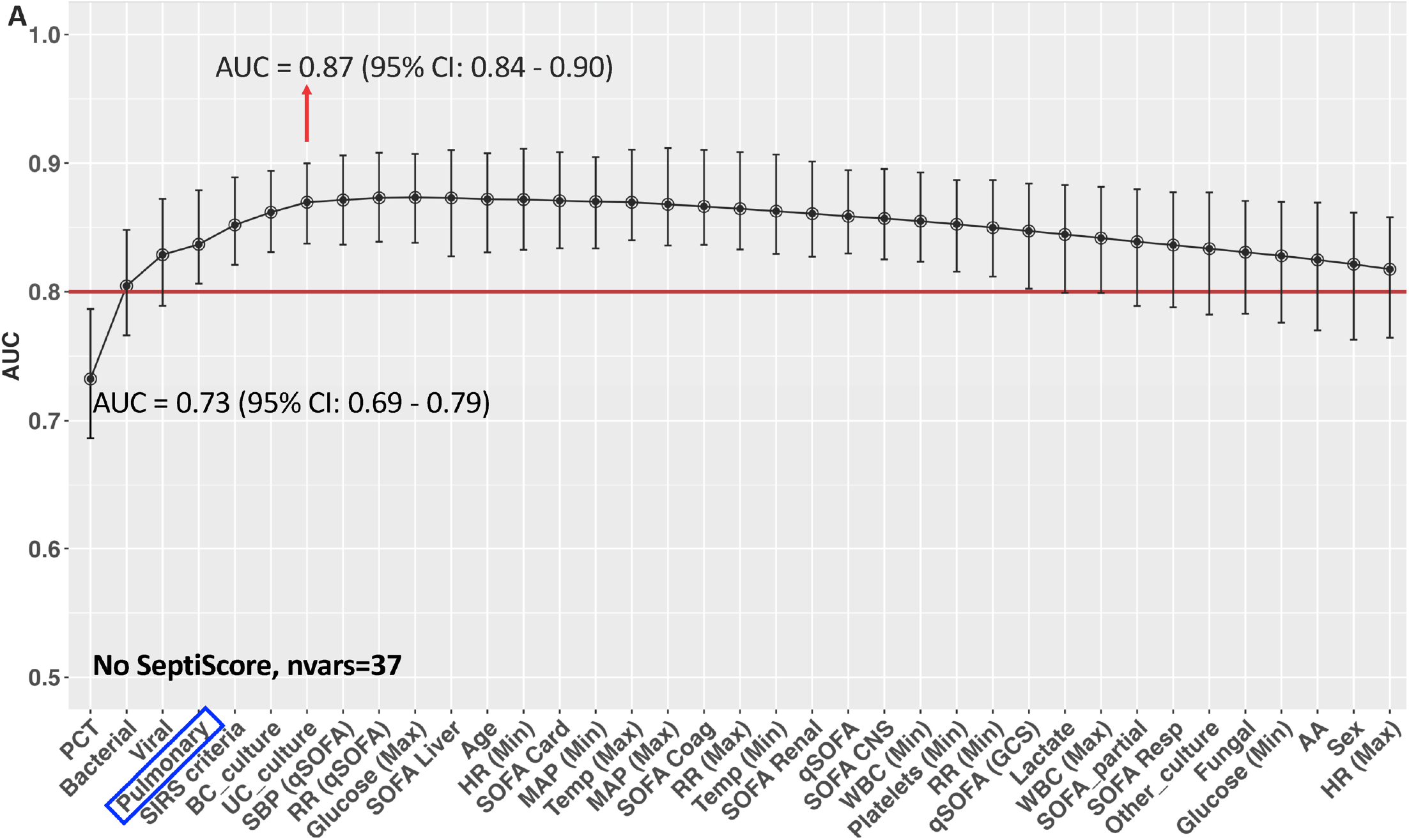
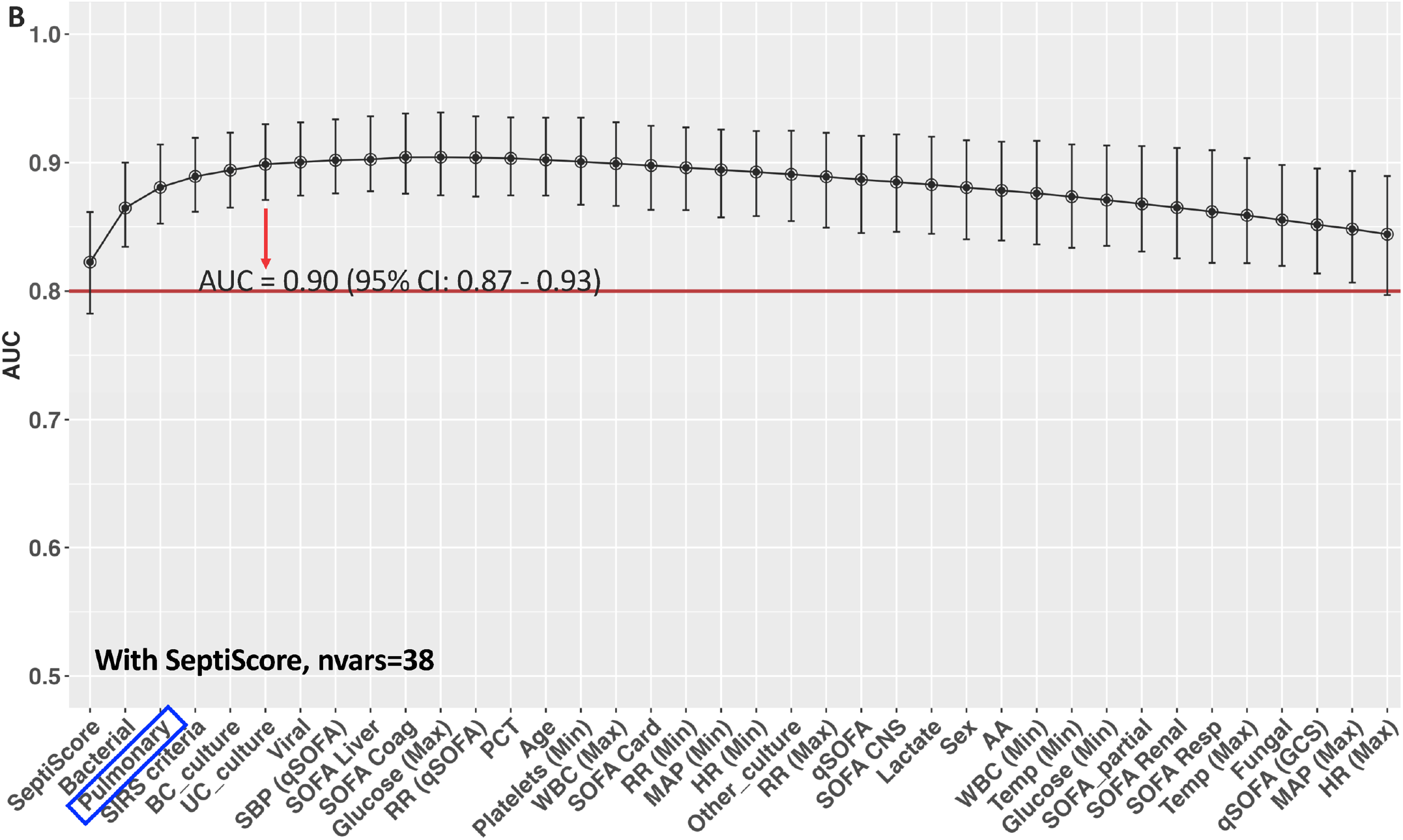
Diagnostic performance based on T1 and T2 parameters, plus microbiological culture results at time T3. Results for Pulmonary site of infection. Data from the 510k cohort. Greedy searches were performed (A) without SeptiCyte RAPID results, and (B) with SeptiCyte RAPID results. Abbreviations: blood culture (BC), urine culture (UC), systolic blood pressure (SBP). Each dot and whisker represent the mean AUC ± 95% CI for the corresponding variable.

Key variables driving performance included SeptiCyte RAPID, PCT, confirmed bacterial or viral infection, site of infection and positive culture results. After the microbiological culture data from time T3 become available, the additional performance boost by including SeptiCyte RAPID is rather small (0.90 – 0.87 = 0.03 AUC units). However, because treatment for sepsis is such a time-critical problem, the benefit of waiting 1-3 days for culture results is out-weighed by the risk of not treating a patient immediately.

### 3.5. Comparison across different suspected sites of infection

Greedy search analyses were also performed on the 510(k) dataset, after incorporating information on suspected or presumed sites of infection. Within a particular time-frame of analysis (T1, T2 or T3), the AUCs did not vary significantly between suspected infection sites.

### 3.6. Analysis of data from the Andalusia Cohort

We conducted comparable greedy search analyses on the data from the Andalusian cohort. AUCs for the Andalusian cohort at successive time points were comparable to AUCs from the 510k cohort, and similarly increased as more diagnostic information became available over time (see Table 2). As in the 510(k) dataset, AUCs in the Andalusian dataset did not vary significantly by the suspected/presumed site of infection, for any of the time-delimited analyses (T1, T2, or T3).

**Table 1.**
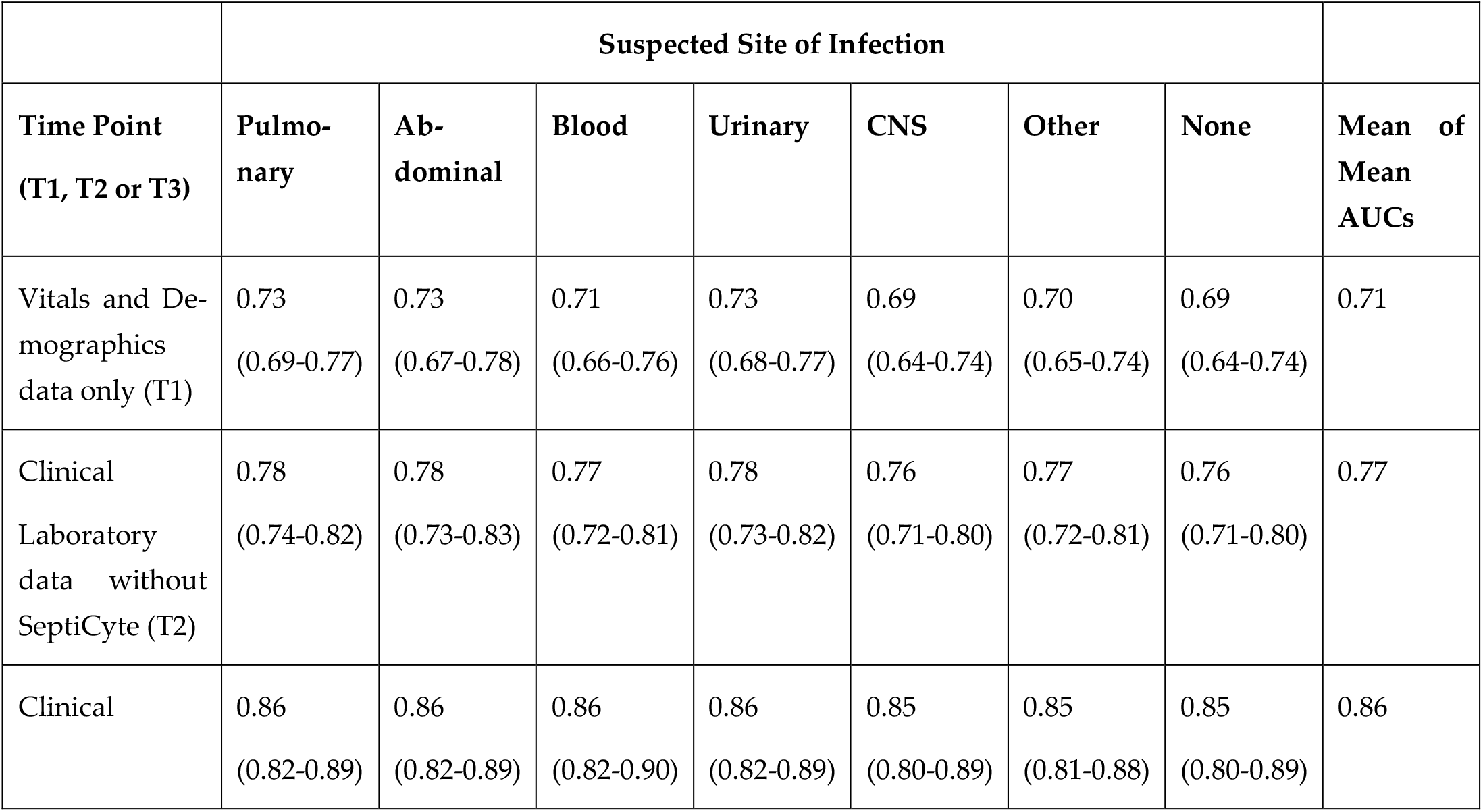
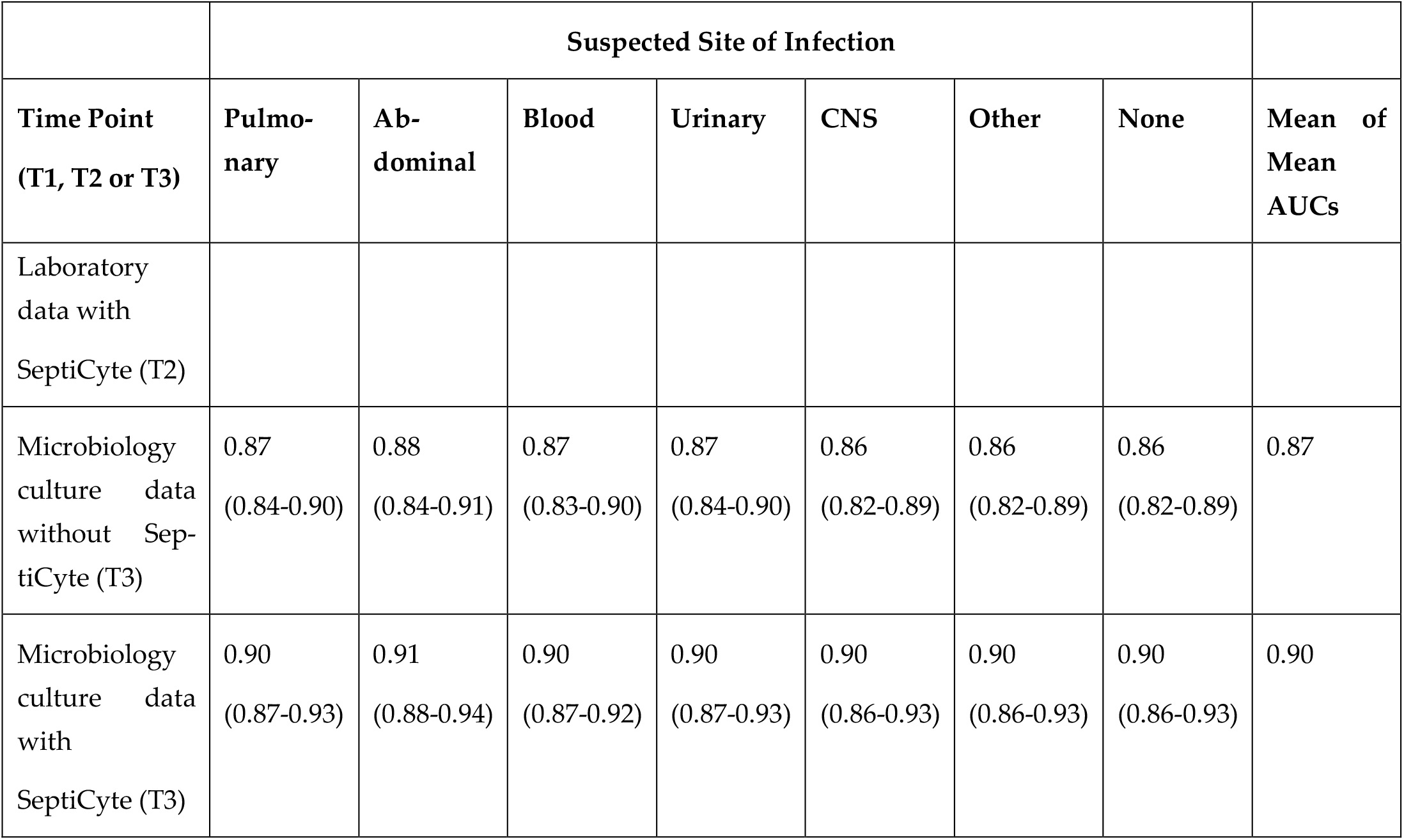
Summary of Performance (mean AUC ± 95% CI) at Three Time Points (T1, T2 or T3), for patients stratified by sites of infection. Data from the 510k cohort.

**Table 2.**
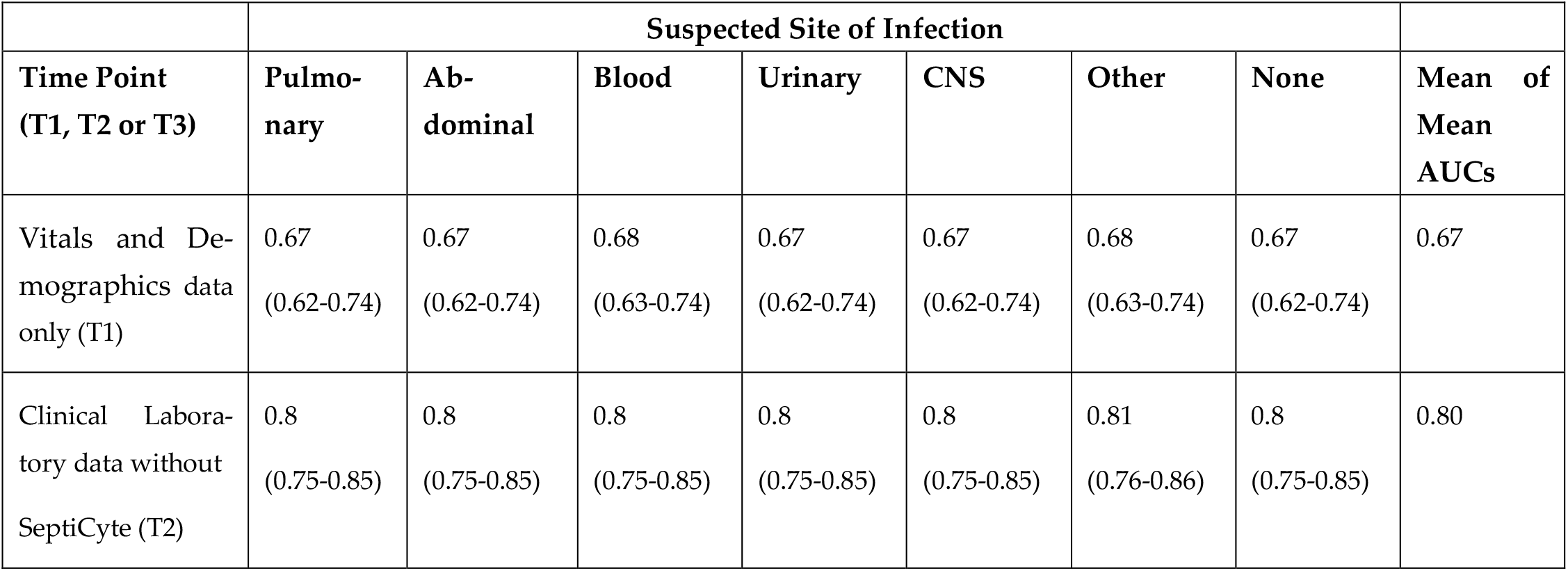
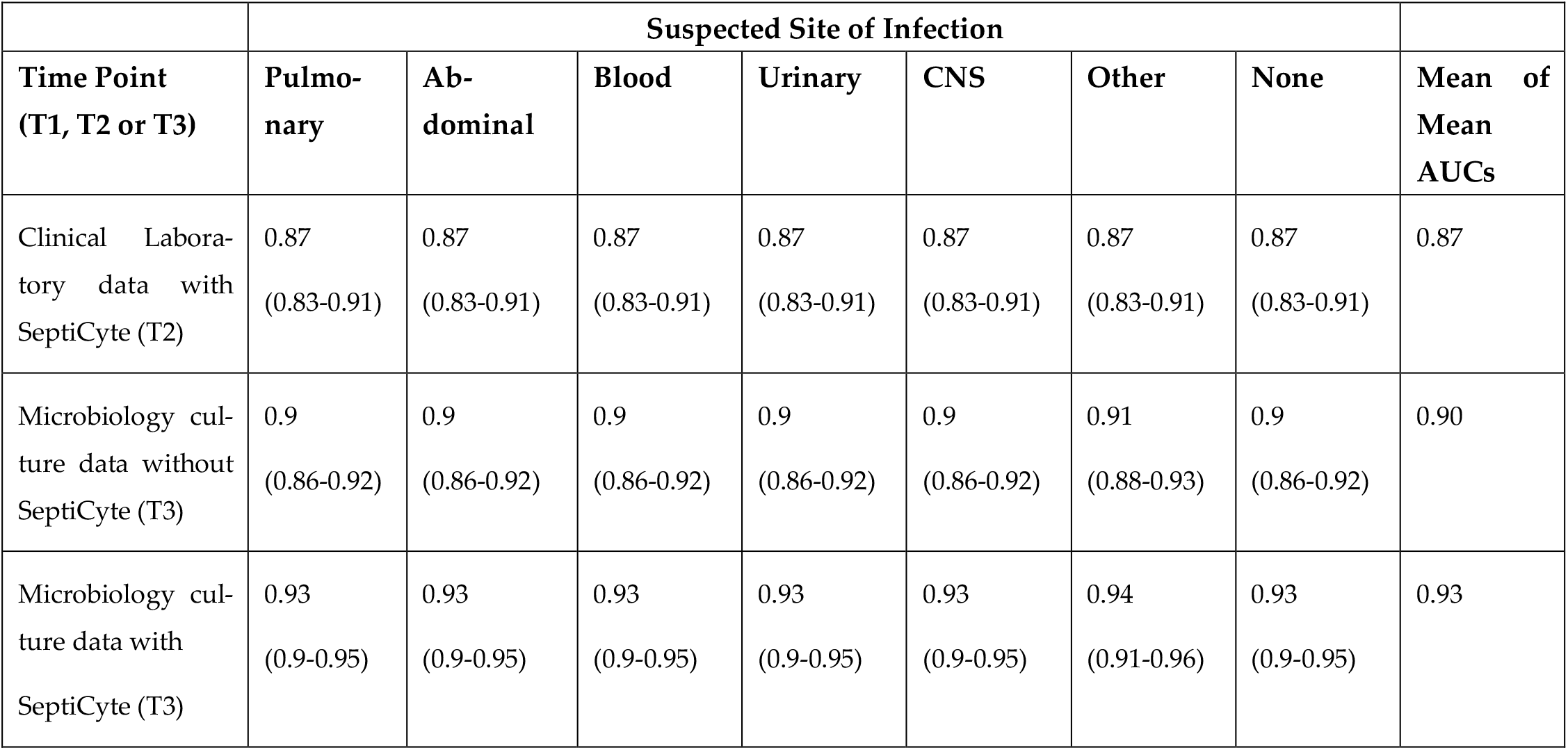
Summary of Performance (mean AUC ± 95% CI) at Three Time Points (T1, T2 or T3), for patients stratified by sites of infection. Data from the Andalusian cohort.

|  | Suspected Site of Infection |  |  |  |  |  |  |  |
| --- | --- | --- | --- | --- | --- | --- | --- | --- |
| Time Point<br>(T1, T2 or T3) | Pulmo-<br>nary | Ab-<br>dominal | Blood | Urinary | CNS | Other | None | Mean of<br>Mean<br>AUCs |
| Vitals and De-<br>mographics data<br>only (T1) | 0.67<br>(0.62-0.74) | 0.67<br>(0.62-0.74) | 0.68<br>(0.63-0.74) | 0.67<br>(0.62-0.74) | 0.67<br>(0.62-0.74) | 0.68<br>(0.63-0.74) | 0.67<br>(0.62-0.74) | 0.67 |
| Clinical Labora-<br>tory data without<br>SeptiCyte (T2) | 0.8<br>(0.75-0.85) | 0.8<br>(0.75-0.85) | 0.8<br>(0.75-0.85) | 0.8<br>(0.75-0.85) | 0.8<br>(0.75-0.85) | 0.81<br>(0.76-0.86) | 0.8<br>(0.75-0.85) | 0.80 |

| Time Point<br>(T1, T2 or T3) | Suspected Site of Infection |  |  |  |  |  |  | Mean of<br>Mean<br>AUCs |
| --- | --- | --- | --- | --- | --- | --- | --- | --- |
|  | Pulmo-<br>nary | Ab-<br>dominal | Blood | Urinary | CNS | Other | None |  |
| Clinical Labora-<br>tory data with<br>SeptiCyte (T2) | 0.87<br>(0.83-0.91) | 0.87<br>(0.83-0.91) | 0.87<br>(0.83-0.91) | 0.87<br>(0.83-0.91) | 0.87<br>(0.83-0.91) | 0.87<br>(0.83-0.91) | 0.87<br>(0.83-0.91) | 0.87 |
| Microbiology cul-<br>ture data without<br>SeptiCyte (T3) | 0.9<br>(0.86-0.92) | 0.9<br>(0.86-0.92) | 0.9<br>(0.86-0.92) | 0.9<br>(0.86-0.92) | 0.9<br>(0.86-0.92) | 0.91<br>(0.88-0.93) | 0.9<br>(0.86-0.92) | 0.90 |
| Microbiology cul-<br>ture data with<br>SeptiCyte (T3) | 0.93<br>(0.9-0.95) | 0.93<br>(0.9-0.95) | 0.93<br>(0.9-0.95) | 0.93<br>(0.9-0.95) | 0.93<br>(0.9-0.95) | 0.94<br>(0.91-0.96) | 0.93<br>(0.9-0.95) | 0.93 |

### 3.7. Comparison of analysis results from the 510k and Andalusian cohorts

It was not possible to formally cross-validate the results from the 510k and Andalusian cohorts, because the variables measured and analyzed in the two cohorts were not exactly the same. Nonetheless, there was a significant overlap of approximately 28 variables (see Supplementary Table S3). While variations in clinical practice from site to site are to be expected, the fact that many of the same variables were tested at different sites, under different study protocols, increases confidence in the generality of the findings described here. Comparing the two sets of analyses, we conclude the following:

1. At time T1 (presentation), regardless of the combination of variables based on patient history, demographics and vital signs, the maximum AUC that could be obtained was around AUC 0.70 ± 0.03.
2. At time T2 (1-3 hours after presentation), the addition of clinical-laboratory variables boosted the AUC to 0.85 ±0.02. SeptiCyte RAPID test results became available at this time, and the SeptiScore had the greatest positive impact on AUC of all variables available at that time.
3. At time T3 (1–3 days after presentation), yet another performance gain was achieved through incorporation of positive microbiological culture results and pathogen identification, boosting the AUC further to 0.88±0.02 without SeptiCyte RAPID, or to AUC 0.90-0.93 with SeptiCyte RAPID. While unsurprising, the microbiological culture and pathogen ID data become available only after the critical early treatment window (T2, 1–3 hours after presentation), when timely initiation of appropriate sepsis therapy is most consequential.
4. In logistic regression, the performance benefit obtained by combining variables typically plateaus after approximately 4–7 variables, after which model performance may begin to decline. This well-recognized phenomenon likely reflects several contributing factors, including sensitivity of the logistic regression model to parameter estimates near the decision boundary, incorporation of noise from variables weakly associated with outcomes, overfitting arising from fitting an increasing number of parameters to a fixed dataset, and redundancy introduced by highly correlated predictor variables [20] (Hastie et al., 2009).

### 3.8. Analysis of potential Incorporation Bias

The present analysis suffers to some extent from the possibility of incorporation bias, because some of the variables used in performance calculations were also used in the reference adjudication process. We have addressed this concern in the following way. A logistic regression model using seven informative variables, identified by greedy feature selection (SeptiCyte RAPID, PCT, Temperature (max), RR (max), MAP (max), age, HR (min)), yielded an AUC of 0.85 (95% CI: 0.80-0.89). A model incorporating only SeptiCyte RAPID and PCT, variables not used by the RPD panel and therefore not subject to incorporation bias, yielded an AUC of 0.83 (95% CI: 0.78-0.87) (See Supplementary Figure S6 and Table S9). Therefore, the majority of the model’s discriminative performance was preserved using variables independent of the RPD. It appears that the variables Temperature (max), RR (max), MAP (max), age, HR (min) together contribute only +0.02 unit to the overall AUC performance measure. A model excluding just SeptiCyte RAPID yielded an AUC of 0.75 (95% CI: 0.70-0.79). Exclusion of Temperature (max) produced the largest reduction in AUC (Δ AUC = −0.01), while exclusion of the remaining variables resulted in smaller changes (Δ AUC < -0.01). No single variable subject to incorporation bias accounted for more than 0.01 AUC unit. We conclude that, in our analysis, incorporation bias exerts a relatively small effect on AUC-based performance estimates.

## 4. Discussion

We have attempted to quantify the diagnostic value of data collected at different times during a patient’s transit through hospital, for diagnosing sepsis vs. SIRS. Data from two independent study cohorts (510k, Andalusia) were examined, with similar results. The 510k cohort consisted of 419 patients, and the Andalusian cohort consisted of 353 patients. Both cohorts were composed of critically ill patients in the ICU, and retrospectively diagnosed with either sepsis or SIRS. Although it was not possible to pool the data from the two cohorts (because not all variables were in common), similar results were obtained in the two cases, lending support to the general approach proposed here.

Diagnostic variables were stratified by their temporal availability to represent the evolving clinical picture within the first three days from hospital admission to ICU care. Three timepoints were considered: T1, immediately upon presentation; T2, within 1-3 hours when clinical-laboratory results become available; and T3, within 1-3 days when microbiological culture results become available. The analysis revealed that diagnostic performance, measured by AUC, increased progressively from ∼0.7 using clinical variables to ∼0.9 when incorporating all variables including microbiological culture and SeptiCyte RAPID results. The greatest single-time-point gain in AUC (from ∼0.7 to ∼0.85) occurred with the inclusion of SeptiCyte RAPID at time T2. Additional clinical and laboratory measures did not add much more diagnostic value, until time T3 when blood culture results became available, which boosted the AUC performance up to ∼0.9.

This study therefore demonstrates that diagnostic performance at time T1 is limited to AUC ∼0.7 using just the data available at presentation (patient history, demographics, vital signs). However, an efficient diagnostic workup at time T2 (1-3 hours after presentation) can substantially improve the accuracy of identifying sepsis in critically ill patients suspected of sepsis. Specific clinical-laboratory variables available at T2 can be used to accurately identify patients with sepsis in lieu of traditional microbiological culture results. We argue that in most patients suspected of sepsis, the results available at T2 are accurate and timely enough to make informed treatment and downstream diagnostic decisions.

### 4.1. Upon Presentation (Time T1)

At presentation (time T1) a limited set of readily measured variables —including vital signs such as maximum temperature, mean arterial pressure (MAP), respiratory rate, and suspected site of infection—provided modest discriminatory power (AUC = 0.69–0.74). This modest discriminatory power is similar to that previously reported in the literature when using SOFA (0.74) and qSOFA (0.73) variables [21] (Rincon et al., 2023). These findings, although limited in terms of AUC, reinforce the importance of prompt clinical assessment and vital sign interpretation, which remain cornerstones of early sepsis recognition [5] (Evans et al., 2021). Interestingly, respiratory rate emerged as a key variable contributing to AUC except in respiratory infections, reflecting its general association with systemic illness rather than site-specific infection. These variables overlap substantially with components of early warning scores such as SIRS, NEWS2, MEWS and qSOFA [22] (Hincapie-Osorno et al., 2024), though qSOFA itself showed limited predictive enhancement in this cohort, aligning with recent critiques of its diagnostic utility [23] (Duncan et al., 2021).

### 4.2. Assessment with Clinical-Laboratory Tests (Time T2)

At time T2 (1-3 hours), clinical-laboratory test results became available and can be added to the diagnostic workup. Diagnostic accuracy improved accordingly (AUC ∼ 0.77). When SeptiCyte RAPID results were included, performance increased to AUC ∼0.85, demonstrating its superior discriminatory performance compared to procalcitonin (PCT) and lactate. PCT, while commonly used, neither outperformed nor acted synergistically with SeptiCyte RAPID, underscoring the latter’s value as an independent biomarker of host response to sepsis, rather than just the presence of a bacterial infection. These findings are consistent with prior studies showing the diagnostic challenges posed by overlapping inflammatory responses and the need for molecular host-response assays to reduce diagnostic uncertainty [4,24] (Lengquist et al., 2024; Roger et al., 2023).

### 4.3. Microbiology Assessment (Time T3)

By 1–3 days post-presentation (Time T3), the incorporation of microbiological culture results further increased AUC to ∼0.9, particularly when combined with the SeptiCyte RAPID scores. Culture results added approximately 0.03-0.04 AUC units of diagnostic gain when SeptiCyte data were already available, reflecting the limited incremental value of delayed microbiological confirmation in guiding early therapy. This suggests that rapid, molecular-based diagnostics may provide clinicians with most of the actionable information needed within the first few hours, a timeframe critical for meeting sepsis management guidelines [25] (Rhee et al., 2021).

### 4.4. Study Limitations

There are several limitations to this study. First, the datasets, though multicentric and demographically diverse, included only ICU patients and may not generalize to non-ICU populations. Nonetheless, the consistently high performance of SeptiCyte RAPID across infection sites supports its robustness as an adjunct to clinical judgment. A second limitation is that clinical-laboratory variables were not consistently collected across the two cohorts, making it challenging to determine commonality of variables that contributed to diagnostic AUC. More extensive studies using more heterogenous patient populations and common variables may be required to confirm the results presented here. Third, this retrospective analysis is subject to some degree of incorporation bias, because a number of variables evaluated for performance were also used by the physician adjudication panel to establish the reference standard. This overlap likely inflates the apparent performance of models incorporating these variables. Incorporation bias is a well-recognized limitation in retrospective studies, particularly those relying on physician adjudication [26–28] Our study design accounted for this by specifically blinding the RPD panel members to SeptiCyte RAPID and PCT, which therefore are not susceptible to this particular source of bias. Our further analyses (Supplemental Figure S4, Table S9) suggested that the inflationary effect of incorporation bias in this study was limited to only about +0.02 AUC units.

### 4.5. Clinical Utility Context

Implementing time-based diagnostic frameworks that integrate rapid molecular tests (pathogen- and host response –based) may optimize both antimicrobial stewardship and patient outcomes. Ultimately, even with complete data, diagnostic accuracy reached a maximum AUC around 0.90, emphasizing the intrinsic complexity of sepsis diagnosis [29] (Lopansri et al., 2019), lack of a true gold standard for sepsis diagnosis [30] (McHugh et al., 2019) and the continuing need for integrated, multimodal approaches [31] (Vincent, 2016).

The results of this analysis align with calls to refine sepsis bundles to balance speed and diagnostic precision [5,32-34] (Evans et al., 2021, Bion et al., 2022, Klompas et al., 2023, Rhee et al., 2023). That is, not all patients suspected of sepsis need sepsis bundles within certain timeframes and that flexibility with respect to timing and bundle use / compliance leads to better outcomes. Therefore, rather than rigid bundles for sepsis, it has been recommended that clinicians focus on a “time-limited course of rapid diagnostic investigation” (Surviving Sepsis Campaign, 2021 Guidelines Adults) or an emphasis on “diagnostic strategies and outcomes” [34] (Rhee et al., 2023).

## 5. Conclusions

We have shown that SeptiCyte RAPID provides valuable information as part of a diagnostic workup and within a limited timeframe. Its use, in combination with other parameters and clinical judgement, may lead to more judicious use of antibiotics, fewer missed diagnoses of “sepsis mimics” and appropriate implementation of sepsis bundles in patients suspected of sepsis that actually have a higher likelihood of sepsis.

## Supplementary Materials

The following supporting information is provided. Table S1. List of Variables and Missing Data Proportions in the 510k cohort. Table S2. List of Variables and Missing Data Proportions in the Andalusian cohort. Table S3. Overlap of Variables Between Analyses of 510k and Andalusian Cohorts. Figure S1 (A-F). Greedy search plots showing performance (AUC) of various clinical variables available at time T1 (presentation), for various initially suspected sites of infection. Figure S2 (A-F). Greedy search plots showing performance (AUC) of various clinical laboratory variables available at time T2 (within 1-3 hours), for various initially suspected sites of infection. Figure S3 (A-F). Greedy search plots showing performance (AUC) of various clinical variables available at Time T3 (within 1-3 days), for various initially suspected sites of infection. Table S4. Greedy Search Results for the Andalusian Cohort, at Time T1 (Presentation). Table S5. Greedy Search Results for the Andalusian Cohort at Time T2 (1-3 Hours Post-Presentation) Without SeptiCyte RAPID. Table S6. Greedy Search Results for the Andalusian Cohort at Time T2 (1-3 Hours Post-Presentation) With SeptiCyte RAPID. Table S7. Greedy Search Results for the Andalusian Cohort at Time T3 (1-3 Days Post-Presentation) Without SeptiCyte RAPID. Table S8. Greedy Search Results for the Andalusian Cohort at Time T3 (1-3 Days Post-Presentation) With SeptiCyte RAPID. Figure S4. Investigation of potential incorporation bias, by comparative ROC curve analysis. Table S9. Relative magnitude of contributions of logistic regression input variables, arranged in decreasing order of impact upon output AUC.

## Author Contributions

**Conceptualization**, Krupa Navalkar, Dayle Sampson, Thomas Yager and Richard Brandon; **Data curation**, Thomas Yager and Silvia Cermelli; **Formal analysis**, Krupa Navalkar, Dayle Sampson, Thomas Yager and Richard Brandon; **Funding acquisition**, José Garnacho-Montero and María Cantón-Bulnes; **Investigation**, Krupa Navalkar and Thomas Yager; **Methodology**, Krupa Navalkar, Thomas Yager, Roy Davis and Richard Brandon; **Project administration**, José Garnacho-Montero, María Cantón-Bulnes, Russell Miller III, John Burke and Richard Brandon; **Software**, Krupa Navalkar; **Supervision**, José Garnacho-Montero, María Cantón-Bulnes, Thomas Yager, Roy Davis, Silvia Cermelli and Richard Brandon; **Validation**, José Garnacho-Montero, María Cantón-Bulnes, José Luis García-Garmendia, Ángel Estella, Adela Fernández-Galilea, Isidro Blanco, Maria Antonia Estecha-Foncea, Marina Gordillo-Resina, Jorge Gómez, Juan Jesús Pineda-Capitán, Carmen Fernández, Ana Ortega, Rosario Villar, Juan Mora-Ordóñez, Sara González-Soto, Antonio Gutierrez-Pizarraya, Robert Balk, Russell Miller III, John Burke, Gourang Patel, Jorge Parada, Marcus Schultz, Brendon Scicluna, Emily Blodget, Santhi Kumar and Dayle Sampson; **Visualization**, Krupa Navalkar; **Writing – original draft**, Thomas Yager and Richard Brandon; **Writing – review & editing**, Krupa Navalkar, Robert Balk, Gourang Patel, Jorge Parada, Marcus Schultz, Dayle Sampson, Thomas Yager, Roy Davis, Silvia Cermelli and Richard Brandon.

## Institutional Review Board Statement

The studies were conducted in accordance with the Declaration of Helsinki and approved by Institutional Review Boards (see below).

The primary cohort was drawn from retrospective (NCT01905033 and NCT02127502; clinicaltrials.gov (accessed on 17 February 2024)) and prospective trials (NCT05469048; clinicaltrials.gov (first submitted 15 July 2022, accessed on 17 February 2024)). The studies were called MARS, VENUS and NEPTUNE. Ethics approval for the MARS trial was given by the Medical Ethics Committee of the Amsterdam Medical Center (approval 156 June 2010, # 10-056C). Ethics approvals for the VENUS trial were given by the relevant Institutional Review Boards as follows: Intermountain Medical Center/Latter Day Saints Hospital (approval, 21 February 2016, # 1024931); Johns Hopkins Hospital (approval 28 January 2016, # IRB00087839); Rush University Medical Center (approval 11 March 2016, # 15111104-IRB01); Loyola University Medical Center (approval, 10 March 2016, # 208291); Northwell Healthcare (approval 1 April 2016, #16-02-42-03). Ethics approvals for the NEPTUNE trial were given by the relevant Institutional Review Boards as follows: Emory University (approval 4 December 2019, # IRB00115400); Grady Memorial Hospital (approval, 14 January 2020, # 00-115400); Rush University Medical Center (approval, 16 January 2020, # 19101603-IRB01); University of Southern California Medical Center (approval, 10 February 2020, # HS-19-0884-CR001). The secondary cohort was conducted in seven ICUs in Andalusia (Spain) and coordinated by the Virgen Macarena University Hospital in Seville. The study was approved by the Research Ethics Committees of the Virgen Macarena-Virgen Rocio University Hospitals (Certificate of Ethics Approval dated 23 Dec 2021. Trial name SEPT-ANC. Codigo Interno: 2662-N-21.)

## Informed Consent Statement

Informed consent was obtained from all subjects involved in the clinical studies. All methods used in this study were carried out in accordance with the relevant guidelines and regulations. For the secondary cohort blood sample extractions were permitted prior to obtaining written permission, if needed, to enable SeptiCyte RAPID results to be generated as quickly as possible. Written consent from the patient, or next of kin, was obtained within 48 hours of ICU admission and the blood sample was discarded if written consent was not obtained.

## Data Availability Statement

The datasets presented in this article are not readily available for patient privacy and commercial reasons. Requests to access the datasets should be directed to Richard Brandon.

## Acknowledgments

Generative AI was not used for study design, data collection, analysis, or interpretation of data, or for generating data or graphics. For some paragraphs within the manuscript. generative AI was used to explore alternative stylistic phrasings to improve clarity for the reader, once the key conceptual points had been articulated by the authors.

## Abbreviations

The following abbreviations are used in this manuscript:

Ag/Ab: Antigen / Antibody (testing)
AST: Antibiotic Susceptibility Testing
AUC: Area Under the receiver operating characteristic Curve
CNS: Central Nervous System
DBP: Diastolic Blood Pressure
HR: Heart Rate
ICU: Intensive Care Unit
ID: Identification
MAP: Mean Arterial Pressure
MEWS: Modified Early Warning Score
NEWS: National Early Warning Score
OECD: Organisation for Economic Co-operation and Development
PCT: Procalcitonin
PLA2G7: Phospholipase A2 Group 7 (gene)
PLAC8: Placenta-specific 8 (gene)
qSOFA: Quick Sequential Organ Failure Assessment
RPD: Retrospective Physician Diagnosis
RR: Respiratory Rate
RT-qPCR: Reverse Transcriptase quantitative Polymerase Chain Reaction
SBP: Systolic Blood Pressure
SEP-1: Severe Sepsis and Septic Shock Early Management Bundle (USA)
SIRS: Systemic Inflammatory Response Syndrome
SOFA: Sequential Organ Failure Assessment
T max: Maximum body temperature in a 24-hour period
USC: University of Southern California
WBC: White Blood Cell (Count)

## Disclaimer/Publisher’s Note

The statements, opinions and data contained in all publications are solely those of the individual author(s) and contributor(s) and not of MDPI and/or the editor(s). MDPI and/or the editor(s) disclaim responsibility for any injury to people or property resulting from any ideas, methods, instructions or products referred to in the content.

