## Supplemental Material for "Sequential application of time-stratified demographic, vital, clinical-laboratory and microbiology variables for accurate and rapid identification of sepsis"

### SUPPLEMENTARY MATERIALS


Table S1. List of Variables and Missing Data Proportions in the 510k cohort

Table S2. List of Variables and Missing Data Proportions in the Andalusian cohort

Table S3. Overlap of Variables Between Analyses of 510k and Andalusian Cohorts

Figure S1 (A-F). Greedy search plots showing performance (AUC) of various clinical variables available at time T1 (presentation), for various initially suspected sites of infection.

Figure S2 (A-F). Greedy search plots showing performance (AUC) of various clinical laboratory variables available at time T2 (within 1-3 hours), for various initially suspected sites of infection.

Figure S3 (A-F). Greedy search plots showing performance (AUC) of various clinical variables available at Time T3 (within 1-3 days), for various initially suspected sites of infection.

Table S4. Greedy Search Results for the Andalusian Cohort, at Time T1 (Presentation).

Table S5. Greedy Search Results for the Andalusian Cohort at Time T2 (1-3 Hours Post-Presentation) Without SeptiCyte RAPID.

Table S6. Greedy Search Results for the Andalusian Cohort at Time T2 (1-3 Hours Post-Presentation) With SeptiCyte RAPID.

Table S7. Greedy Search Results for the Andalusian Cohort at Time T3 (1-3 Days Post-Presentation) Without SeptiCyte RAPID.

Table S8. Greedy Search Results for the Andalusian Cohort at Time T3 (1-3 Days Post-Presentation) With SeptiCyte RAPID.

Figure S4. Investigation of potential incorporation bias, by comparative ROC curve analysis.

Table S9. Relative magnitude of contributions of logistic regression input variables, arranged in decreasing order of impact upon output AUC.

**Supplementary Table S1. List of Variables and Missing Data Proportions in the 510k Cohort**

| No. | Variable | Unit | No. of missing values | | |
|---|---|---|---|---|---|
| | | | Sepsis | SIRS | Total |
| | | | N = 176 | N = 243 | N = 419 |
| 1 | SeptiScore | Range 1-15 | 0 | 0 | 0 |
| 2 | PCT | ng/ml | 27 | 45 | 72 |
| 3 | Lactate | mmol/L | 47 | 93 | 140 |
| 4 | Glucose (Max) | mg/dL | 8 | 21 | 29 |
| 5 | Glucose (Min) | mg/dL | 5 | 21 | 26 |
| 6 | Respiratory Rate (Max) | breaths per minute | 53 | 71 | 124 |
| 7 | Respiratory Rate (Min) | breaths per minute | 56 | 73 | 129 |
| 8 | Mean Arterial Pressure (Max) | mmHg | 41 | 54 | 95 |
| 9 | Mean Arterial Pressure (Min) | mmHg | 2 | 7 | 9 |
| 10 | Temperature (Max) | ºC | 19 | 34 | 53 |
| 11 | Temperature (Min) | ºC | 24 | 40 | 64 |
| 12 | Heart Rate (Max) | beats per minute | 0 | 0 | 0 |
| 13 | Heart Rate (Min) | beats per minute | 1 | 1 | 2 |
| 14 | WBC (Max) | x $10^3$ /uL | 7 | 9 | 16 |
| 15 | WBC (Min) | x $10^3$ /uL | 5 | 7 | 12 |
| 16 | Platelets (Min) | x $10^3$ /uL | 52 | 70 | 122 |
| 17 | Age | years | 0 | 0 | 0 |
| 18 | SOFA score | categorical variable | 19 | 27 | 46 |
| 19 | SOFA Cardiovascular component | categorical variable | 22 | 29 | 51 |
| 20 | SOFA Respiratory component | categorical variable | 43 | 73 | 116 |
| 21 | SOFA Liver component | categorical variable | 30 | 63 | 93 |
| 22 | SOFA Coagulation component | categorical variable | 19 | 31 | 50 |
| 23 | SOFA Renal component | categorical variable | 20 | 36 | 56 |
| 24 | SOFA CNS component | categorical variable | 25 | 33 | 58 |
| 25 | qSOFA | categorical variable | 16 | 15 | 31 |

| No. | Variable | Unit | No. of missing values | | |
|---|---|---|---|---|---|
| | | | Sepsis | SIRS | Total |
| | | | N = 176 | N = 243 | N = 419 |
| 26 | RR (max) >=22 (qSOFA) | categorical variable | 53 | 71 | 124 |
| 27 | SBP (min)<=100 (qSOFA) | categorical variable | 96 | 123 | 219 |
| 28 | Sex | categorical variable | 0 | 0 | 0 |
| 29 | African American or not | categorical variable | 0 | 0 | 0 |
| 30 | SIRS criteria | categorical variable | 0 | 0 | 0 |
| 31 | Bacterial pathogen | categorical variable | 0 | 0 | 0 |
| 32 | Viral pathogen | categorical variable | 0 | 0 | 0 |
| 33 | Fungal pathogen | categorical variable | 0 | 0 | 0 |
| 34 | Other culture | categorical variable | 0 | 0 | 0 |
| 35 | Urine (site of infection) | categorical variable | 0 | 0 | 0 |
| 36 | Abdominal (site of infection) | categorical variable | 0 | 0 | 0 |
| 37 | Blood (site of infection) | categorical variable | 1 | 67 | 68 |
| 38 | Central Nervous System (site of infection) | categorical variable | 1 | 67 | 68 |
| 39 | Other (site of infection) | categorical variable | 0 | 0 | 0 |
| 40 | Pulmonary (site of infection) | categorical variable | 1 | 67 | 68 |
| 41 | Blood culture | categorical variable | 0 | 0 | 0 |
| 42 | Urine culture | categorical variable | 0 | 0 | 0 |

**Supplementary Table S2. List of Variables and Missing Data Proportions in the Andalusian Cohort**

| No. | Variable | Unit | No. of missing values | | |
|---|---|---|---|---|---|
| | | | Sepsis | SIRS | Total |
| | | | N = 267 | N = 86 | N=353 |
| 1 | SeptiScore | Range 1-15 | 0 | 0 | 0 |
| 2 | CRP | mg/L | 3 | 1 | 4 |
| 3 | PCT | ng/ml | 4 | 4 | 8 |
| 4 | Lactate | mmol/L | 0 | 0 | 0 |
| 5 | WBC (Leucocytes) | cells/µl | 0 | 0 | 0 |
| 6 | Age | years | 0 | 0 | 0 |
| 7 | Temperature | ºC | 0 | 0 | 0 |
| 8 | Heart Rate | beats per minute | 0 | 0 | 0 |
| 9 | Respiratory Rate | breaths per minute | 0 | 0 | 0 |
| 10 | Mean Arterial Pressure | mmHg | 0 | 0 | 0 |
| 11 | Diastolic Blood Pressure | mmHg | 0 | 0 | 0 |
| 12 | Systolic Blood Pressure | mmHg | 0 | 0 | 0 |
| 13 | Platelets | cells/µl | 0 | 0 | 0 |
| 14 | Creatinine | mg/dL | 0 | 0 | 0 |
| 15 | Neutrophils | % WBC | 0 | 0 | 0 |
| 16 | Lymphocytes | % WBC | 0 | 0 | 0 |
| 17 | Eosinophils | % WBC | 0 | 0 | 0 |
| 18 | Hemoglobin | g/dL | 0 | 0 | 0 |
| 19 | Urea | mg/dL | 9 | 7 | 16 |
| 20 | SOFA (min) at admission | categorical variable | 0 | 0 | 0 |
| 21 | APACHE-II score | categorical variable | 0 | 0 | 0 |
| 22 | Bacterial pathogen identified | categorical variable | 0 | 0 | 0 |
| 23 | Viral pathogen identified | categorical variable | 0 | 0 | 0 |
| 24 | Fungal pathogen identified | categorical variable | 0 | 0 | 0 |
| 25 | Sex | categorical variable | 0 | 0 | 0 |
| 26 | Blood Culture Positive | categorical variable | 0 | 0 | 0 |
| 27 | Other Culture Positive | categorical variable | 0 | 0 | 0 |

| No. | Variable | Unit | No. of missing values | | |
|---|---|---|---|---|---|
| | | | **Sepsis** | SIRS | **Total** |
| | | | **N = 267** | **N = 86** | **N=353** |
| 28 | Urine Culture Positive | categorical variable | 0 | 0 | 0 |
| 29 | Abdominal (site of infection) | categorical variable | 0 | 0 | 0 |
| 30 | Pulmonary (site of infection) | categorical variable | 0 | 0 | 0 |
| 31 | Blood (site of infection) | categorical variable | 0 | 0 | 0 |
| 32 | Urinary (site of infection) | categorical variable | 0 | 0 | 0 |
| 33 | Central Nervous System (site of infection) | categorical variable | 0 | 0 | 0 |
| 34 | Other (site of infection) | categorical variable | 0 | 0 | 0 |
| 35 | SBP<=100 (qSOFA) | categorical variable | 0 | 0 | 0 |
| 36 | RR>=22 (qSOFA) | categorical variable | 0 | 0 | 0 |
| 37 | qSOFA (partial, no GCS) | categorical variable | 0 | 0 | 0 |
| 38 | SIRS criteria | categorical variable | 0 | 0 | 0 |

### Supplementary Table S3. Overlap of Variables Between Analyses of 510k and Andalusian Cohorts

#### 1. Variables in common

| ID # | 510k analysis | Andalusia analysis |
|---|---|---|
| | **Variable** | **Variable** |
| 1 | SeptiScore | SeptiScore |
| 2 | PCT | PCT |
| 3 | Lactate | Lactate |
| 6 | Respiratory Rate (Max) | Respiratory Rate |
| 7 | Respiratory Rate (Min) | |
| 8 | Mean Arterial Pressure (Max) | Mean Arterial Pressure |
| 9 | Mean Arterial Pressure (Min) | |
| 10 | Temperature (Max) | Temperature |
| 11 | Temperature (Min) | |
| 12 | Heart Rate (Max) | Heart Rate |
| 13 | Heart Rate (Min) | |
| 14 | WBC (Max) | WBC (Leucocytes) |
| 15 | WBC (Min) | |
| 16 | Platelets (Min) | Platelets |
| 17 | Age | Age |
| 18 | SOFA score | SOFA (min) at admission |
| 25 | qSOFA | qSOFA (partial, no GCS) |
| 26 | RR (max) >=22 (qSOFA) | RR>=22 (qSOFA) |
| 27 | SBP (min)<=100 (qSOFA) | SBP<=100 (qSOFA) |
| 28 | Sex | Sex |
| 30 | SIRS criteria | SIRS criteria |
| 31 | Bacterial pathogen | Bacterial pathogen |
| 32 | Viral pathogen | Viral pathogen |
| 33 | Fungal pathogen | Fungal pathogen |
| 34 | Other culture positive | Other culture positive |
| 35 | Urinary (site of infection) | Urinary (site of infection) |
| 36 | Abdominal (site of infection) | Abdominal (site of infection) |
| 37 | Blood (site of infection) | Blood (site of infection) |
| 38 | Central Nervous System (site of infection) | Central Nervous System (site of infection) |
| 39 | Other (site of infection) | Other (site of infection) |
| 40 | Pulmonary (site of infection) | Pulmonary (site of infection) |
| 41 | Blood culture positive | Blood culture positive |
| 42 | Urine culture positive | Urine culture positive |

**2. Variables in the Andalusian Analysis, but not in the 510k analysis**

| No. | Variable |
|---|---|
| 1 | CRP |
| 2 | Diastolic Blood Pressure |
| 3 | Systolic Blood Pressure |
| 4 | Creatinine |
| 5 | Neutrophils |
| 6 | Lymphocytes |
| 7 | Eosinophils |
| 8 | Hemoglobin |
| 9 | Urea |
| 10 | APACHE-II score |

**3. Variable in the 510k Analysis, but not in the Andalusian Analysis**

| No. | Variable |
|---|---|
| 1 | Glucose (Max) |
| 2 | Glucose (Min) |
| 3 | SOFA Cardiovascular component |
| 4 | SOFA Respiratory component |
| 5 | SOFA Liver component |
| 6 | SOFA Coagulation component |
| 7 | SOFA Renal component |
| 8 | SOFA CNS component |
| 9 | African American or not |

**Supplementary Figure S1 (A-F).** Greedy search plots showing performance (AUC) of various clinical variables available at time T1 (presentation), for various initially suspected sites of infection. Data from 510k cohort. A: Abdominal, B: Urinary, C: Blood, D: CNS, E: Other, F: Site of Infection not identified.

**A: Abdominal Initial Impression of Site of Infection**

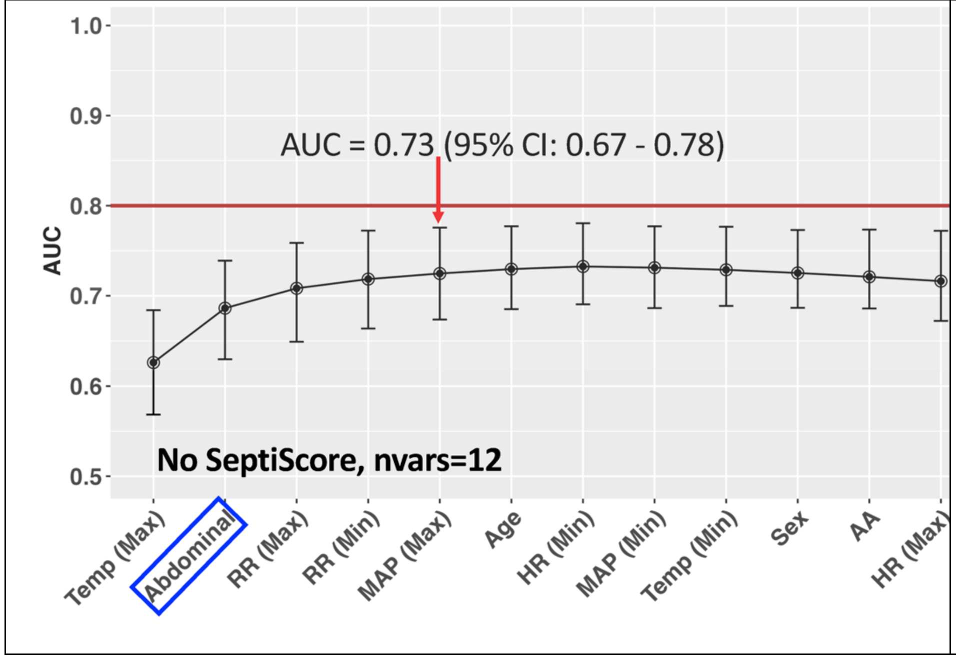


**B: Urinary Initial Impression of Site of Infection**

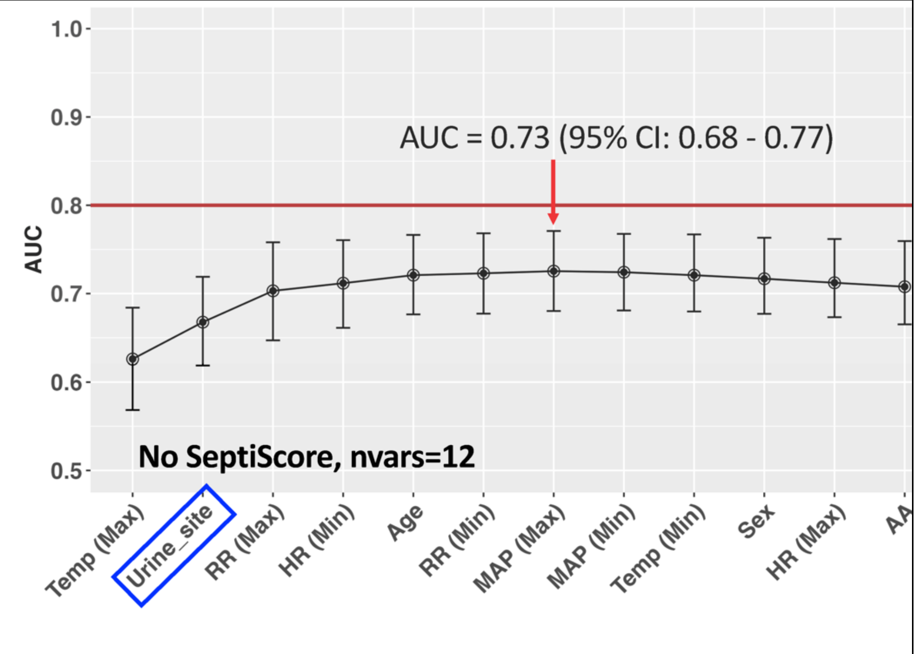

**C: Blood Initial Impression of Site of Infection**

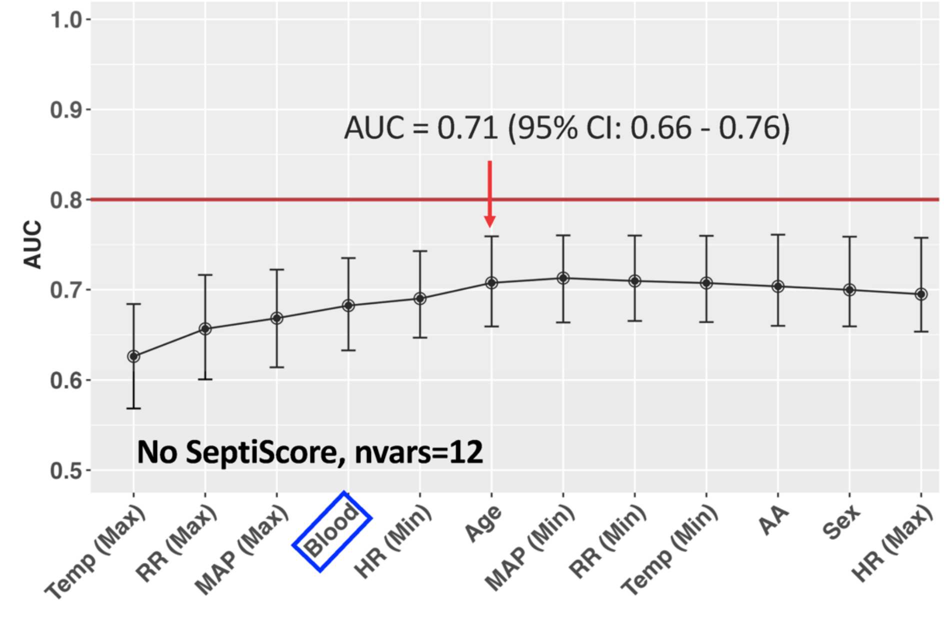


**D: CNS Initial Impression of Site of Infection**

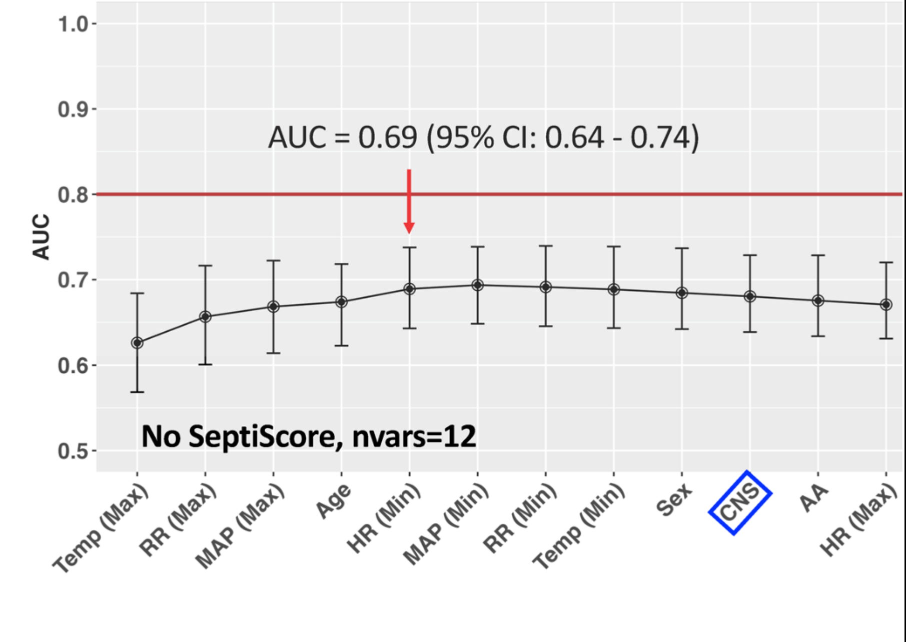

**E: Other Initial Impression of Site of Infection**

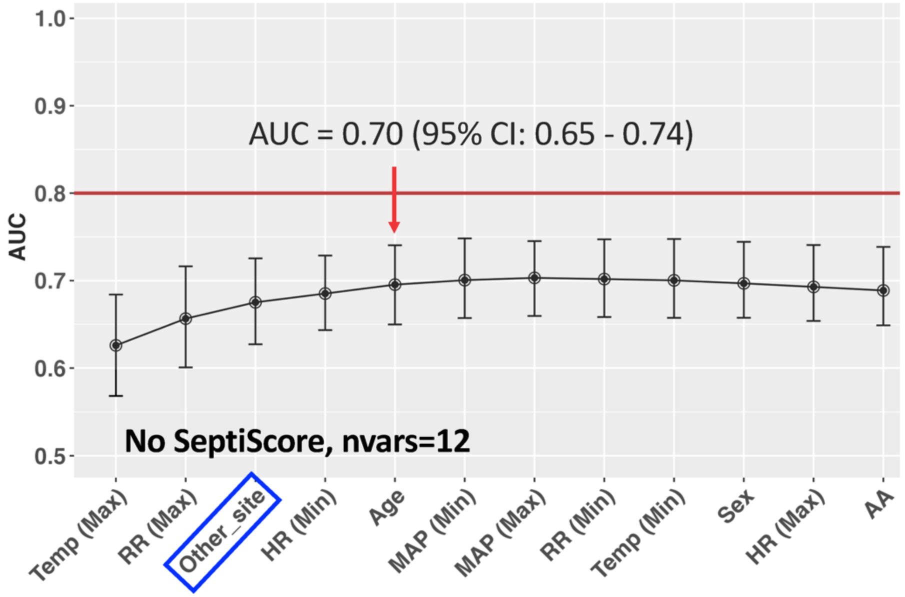


**F: No Initial Site of Infection**

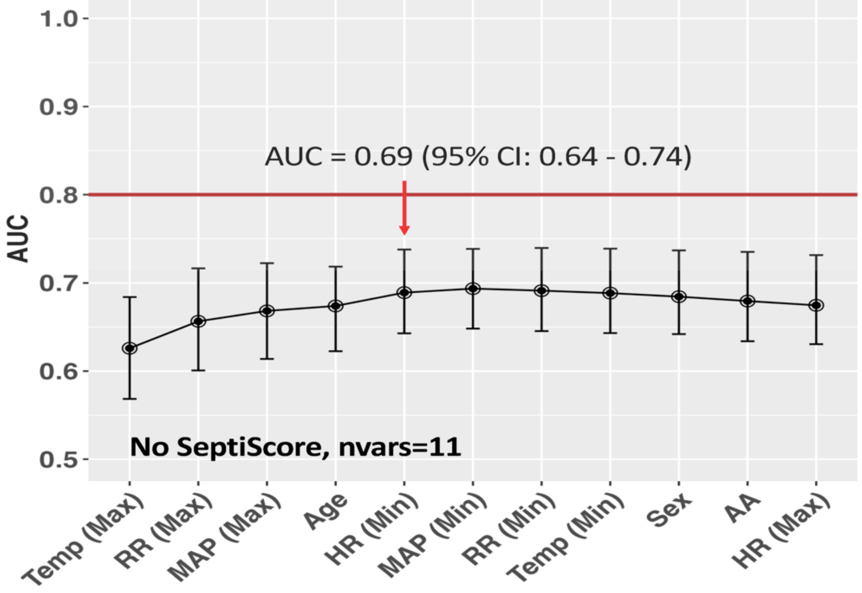

**Supplementary Figure S2 (A-F).** Greedy search plots showing performance (AUC) of various clinical variables available at Time T2 (in the first 1-3 hours) for various sites of infection without (left panel) and with SeptiCyte RAPID (right panel), including A: Abdominal, B: Urinary, C: Blood, D: CNS and E: Other. F: No site of infection identified/ documented at initial clinical impression

**A: Abdominal**

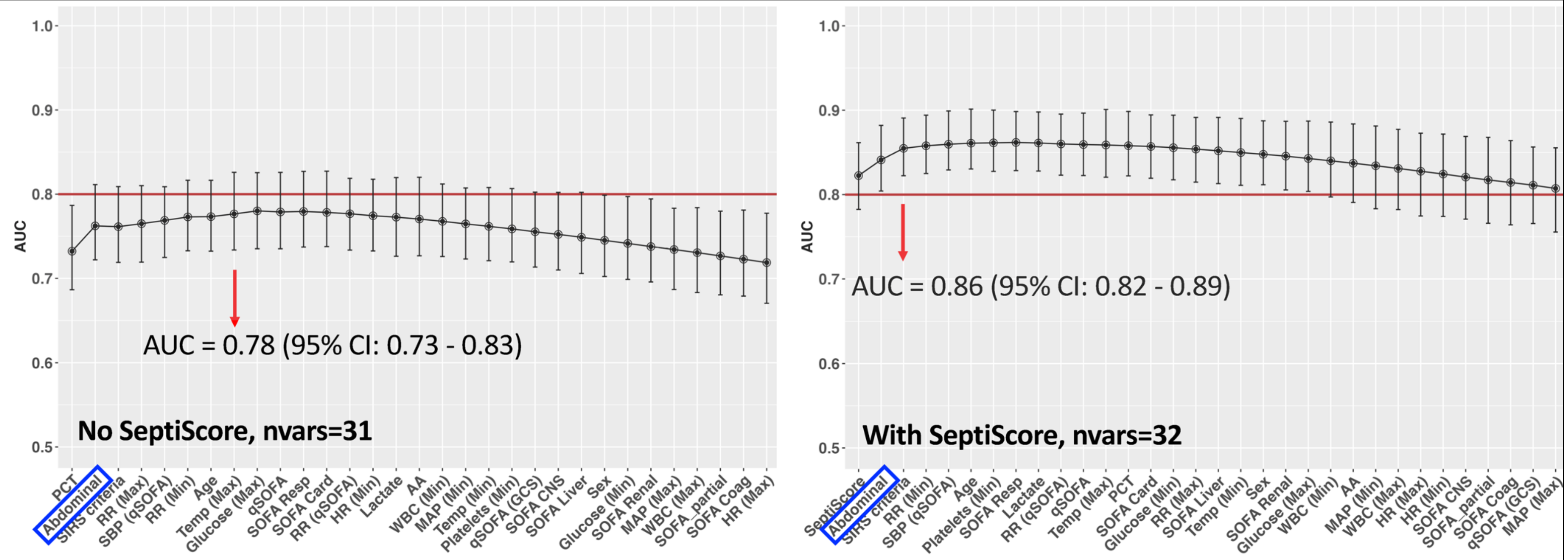

**B: Urinary**

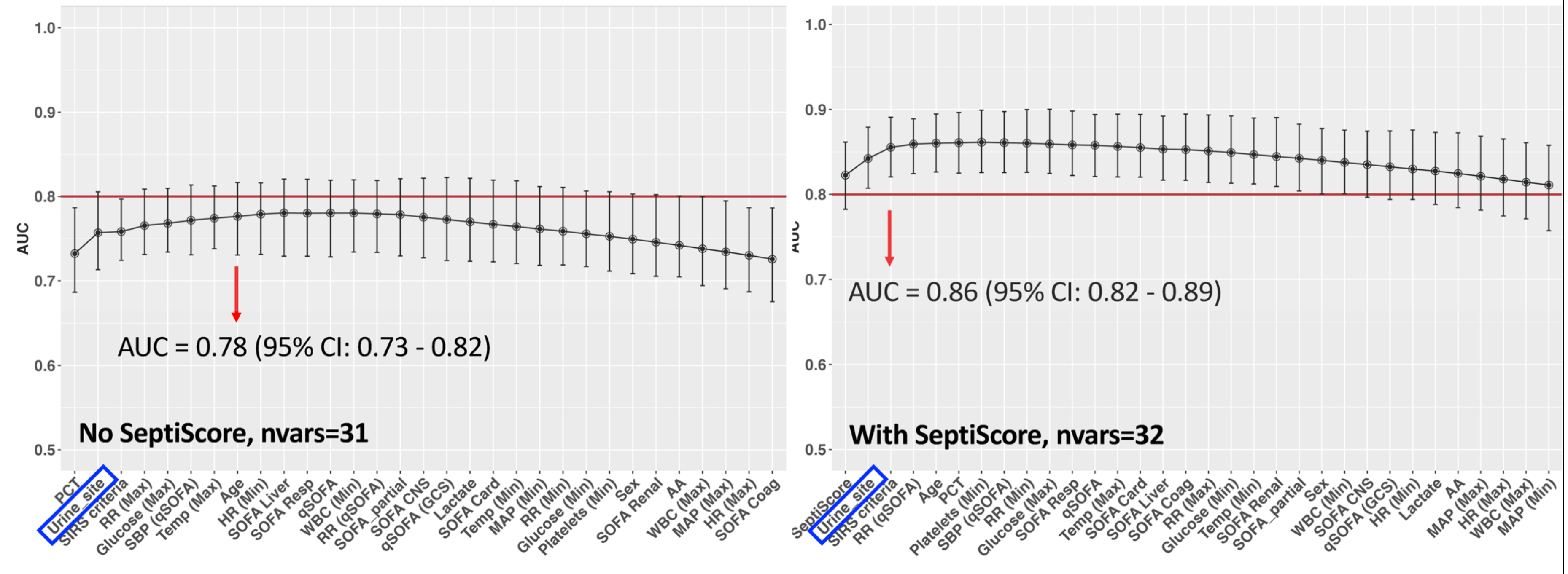
AUC
1.0
0.9
0.8
0.7
0.6
0.5
AUC = 0.78 (95% CI: 0.73 - 0.82)
No SeptiScore, nvars=31
PCT
Urine_site
SIRS criteria
RR (Max)
Glucose (Max)
SBP (qSOFA)
Temp (Max)
Age
HR (Min)
SOFA Liver
SOFA Resp
qSOFA
WBC (Min)
RR (qSOFA)
SOFA_partial
SOFA CNS
qSOFA (GCS)
Lactate
SOFA Card
Temp (Min)
MAP (Min)
RR (Min)
Glucose (Min)
Platelets (Min)
Sex
SOFA Renal
AA
WBC (Max)
MAP (Max)
HR (Max)
SOFA Coag
AUC = 0.86 (95% CI: 0.82 - 0.89)
With SeptiScore, nvars=32
SeptiScore
Urine_site
SIRS criteria
RR (qSOFA)
Age
PCT
Platelets (Min)
SBP (qSOFA)
RR (Min)
Glucose (Max)
SOFA Resp
qSOFA
Temp (Max)
SOFA Card
SOFA Liver
SOFA Coag
RR (Max)
Glucose (Min)
Temp (Min)
SOFA Renal
SOFA_partial
Sex
WBC (Min)
SOFA CNS
qSOFA (GCS)
HR (Min)
Lactate
AA
MAP (Max)
HR (Max)
WBC (Max)
MAP (Min)

C: Blood (Bacteremia)

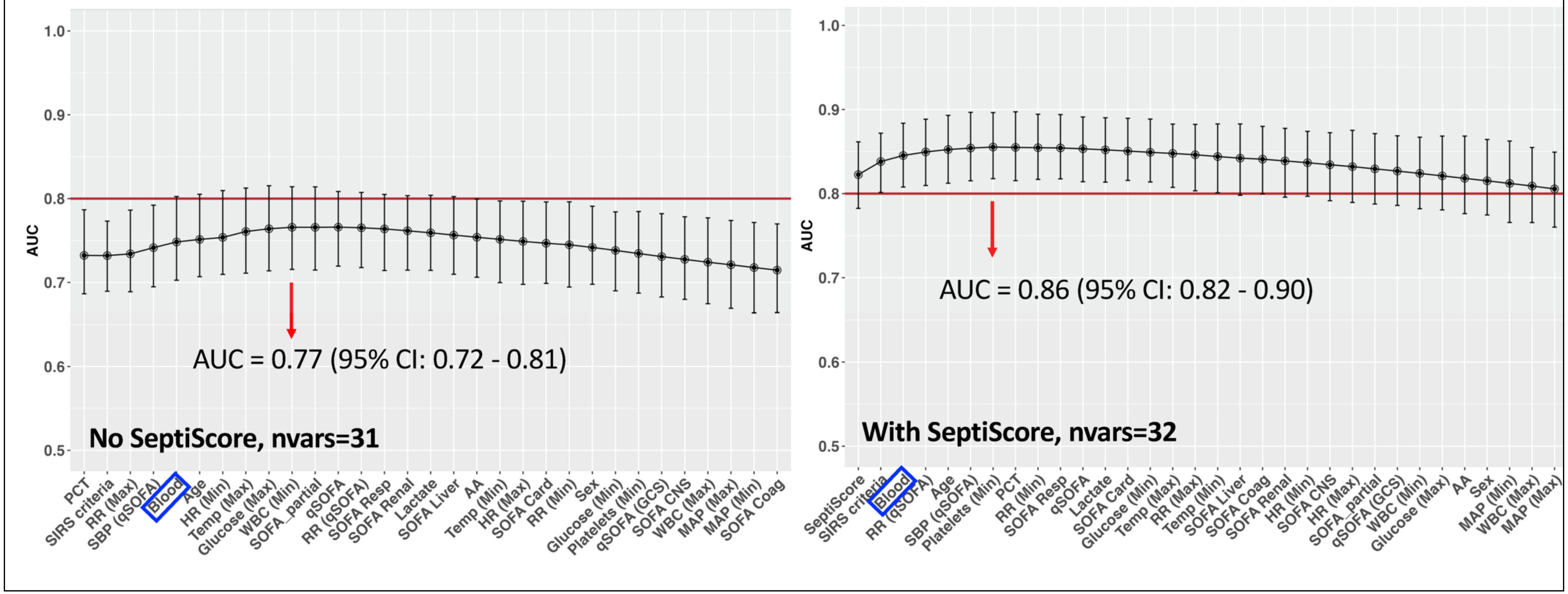

**D: CNS**

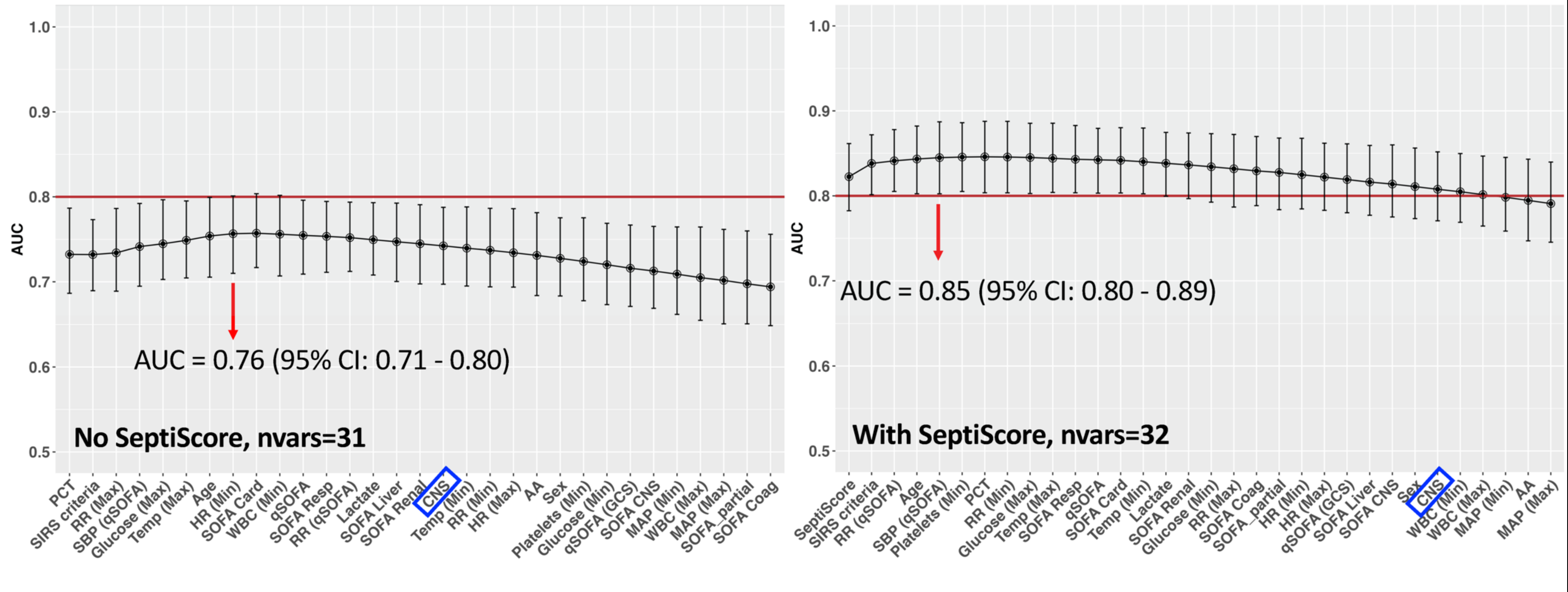
AUC
1.0
0.9
0.8
0.7
0.6
0.5
AUC = 0.76 (95% CI: 0.71 - 0.80)
No SeptiScore, nvars=31
PCT
SIRS criteria
RR (Max)
SBP (qSOFA)
Glucose (Max)
Temp (Max)
Age
HR (Min)
SOFA Card
WBC (Min)
qSOFA
SOFA Resp
RR (qSOFA)
Lactate
SOFA Liver
SOFA Renal
CNS
Temp (Min)
RR (Min)
HR (Max)
AA
Sex
Platelets (Min)
Glucose (Min)
qSOFA (GCS)
SOFA CNS
MAP (Min)
WBC (Max)
MAP (Max)
SOFA_partial
SOFA Coag
AUC = 0.85 (95% CI: 0.80 - 0.89)
With SeptiScore, nvars=32
SeptiScore
SIRS criteria
RR (qSOFA)
Age
SBP (qSOFA)
Platelets (Min)
PCT
RR (Min)
Glucose (Max)
Temp (Max)
SOFA Resp
qSOFA
SOFA Card
Temp (Min)
Lactate
SOFA Renal
Glucose (Min)
RR (Max)
SOFA Coag
SOFA_partial
HR (Min)
HR (Max)
qSOFA (GCS)
SOFA Liver
SOFA CNS
Sex
CNS
WBC (Min)
WBC (Max)
MAP (Min)
AA
MAP (Max)

**E: Other Site of Infection**

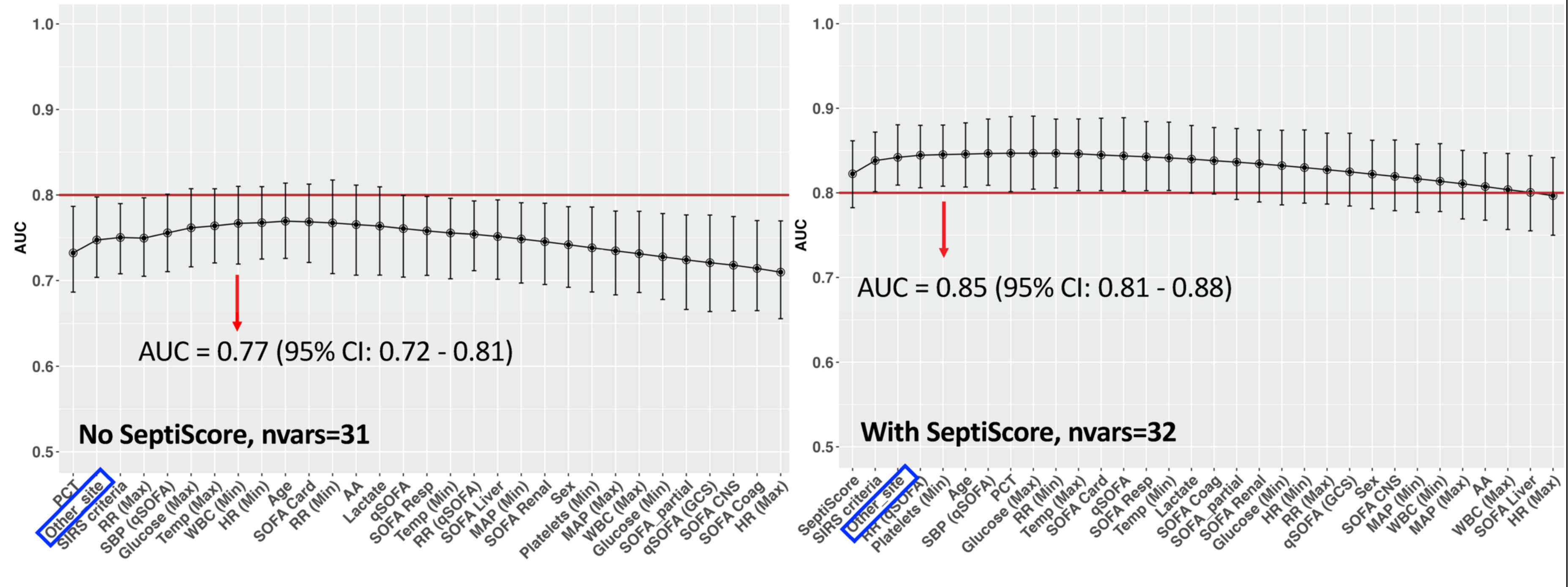

**F: No Site of Infection identified/ documented at initial clinical impression**

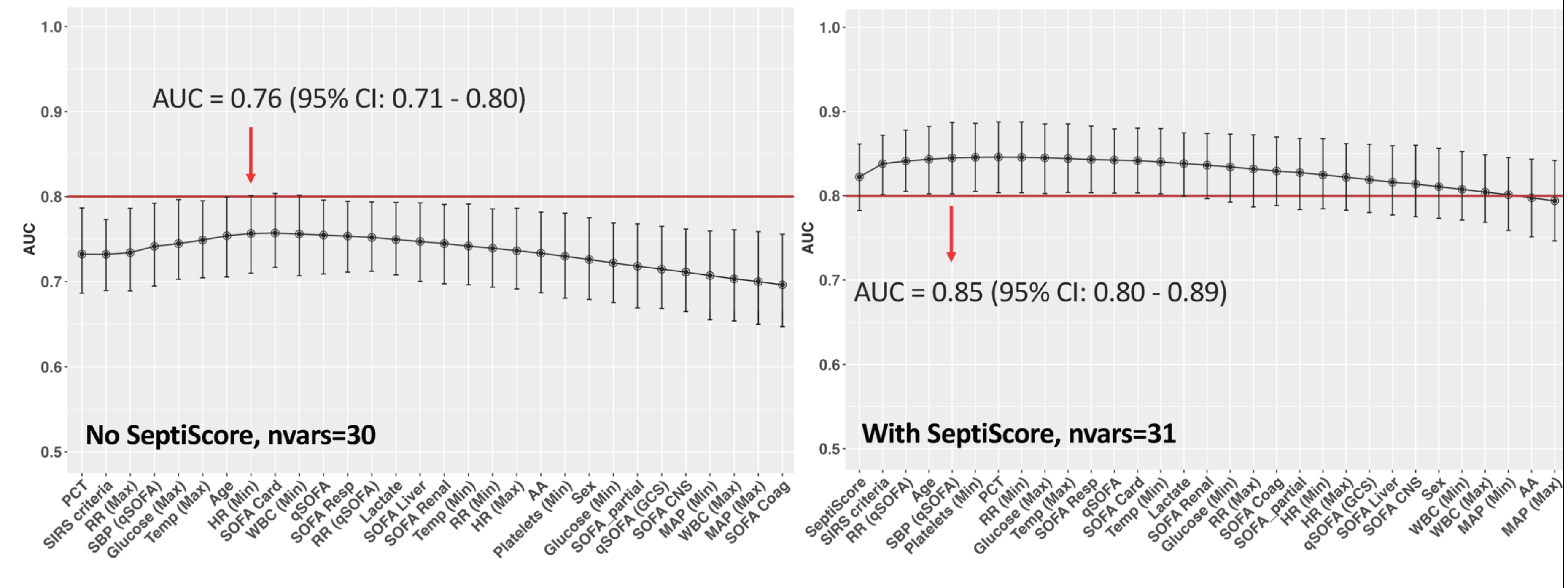

**Supplementary Figure S3 (A-F).** Greedy search plots showing performance (AUC) of various clinical variables available at Time T3 (within 1-3 days) for various sites of infection without (top panel) and with SeptiCyte RAPID (bottom panel), including A: Abdominal, B: Urinary, C: Blood, D: CNS, E: Other and F: No initial impression of site of infection.

**A: Abdominal**

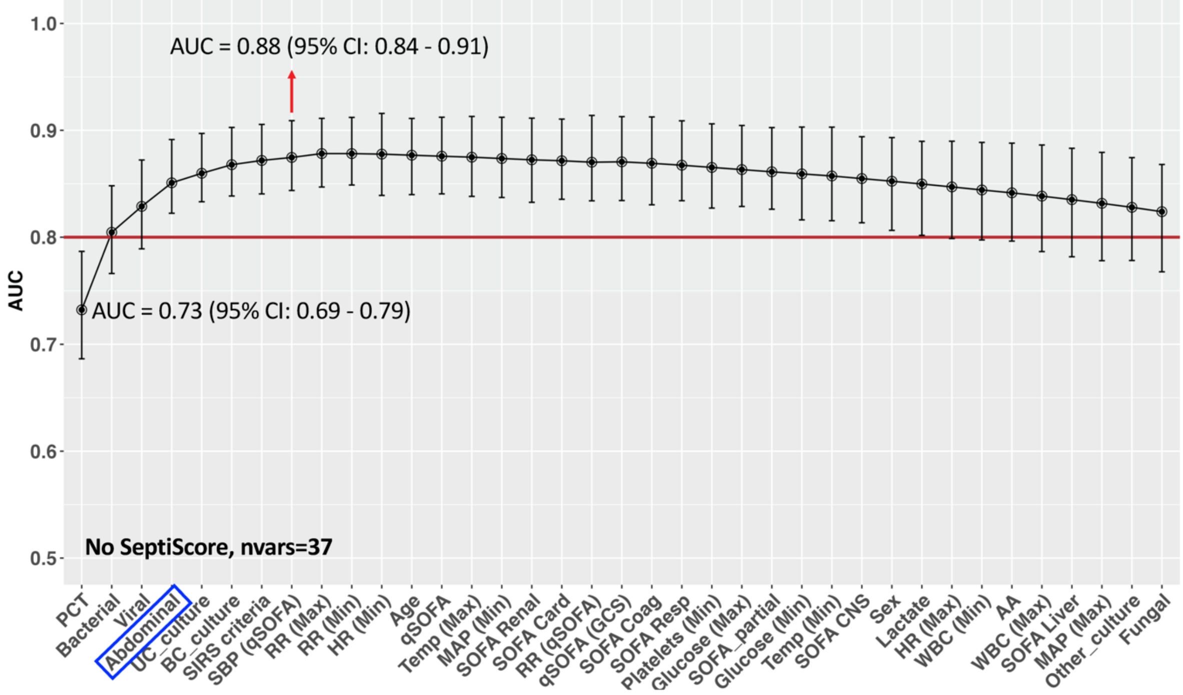


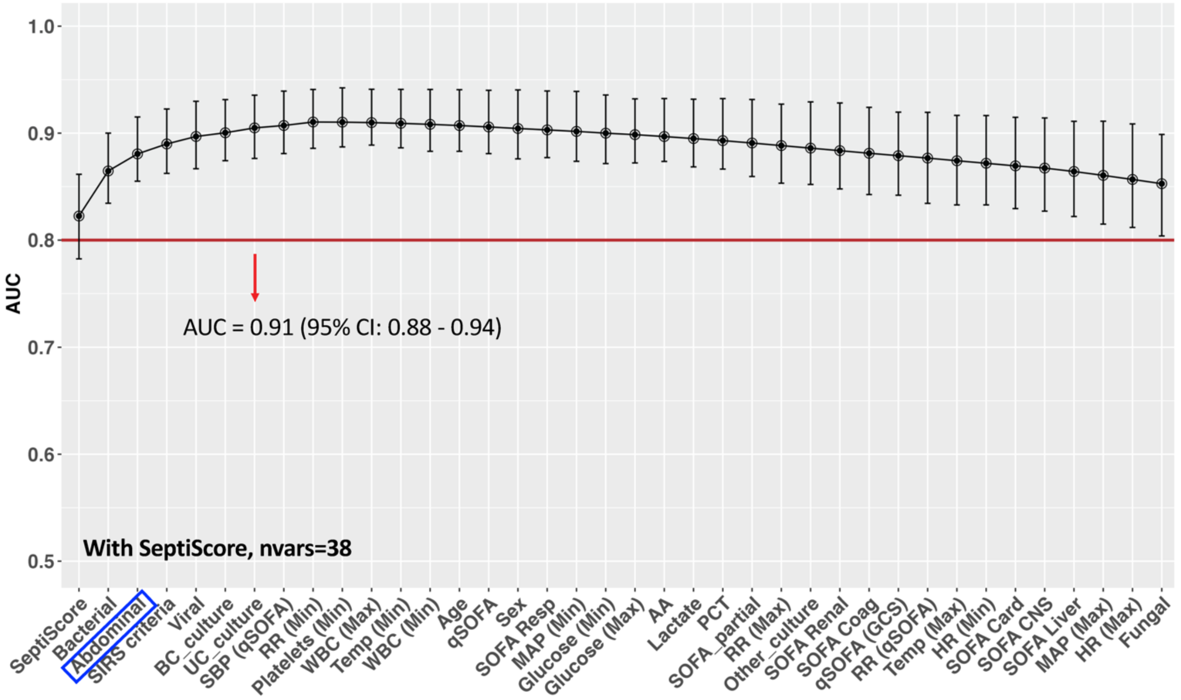

**B: Urinary**

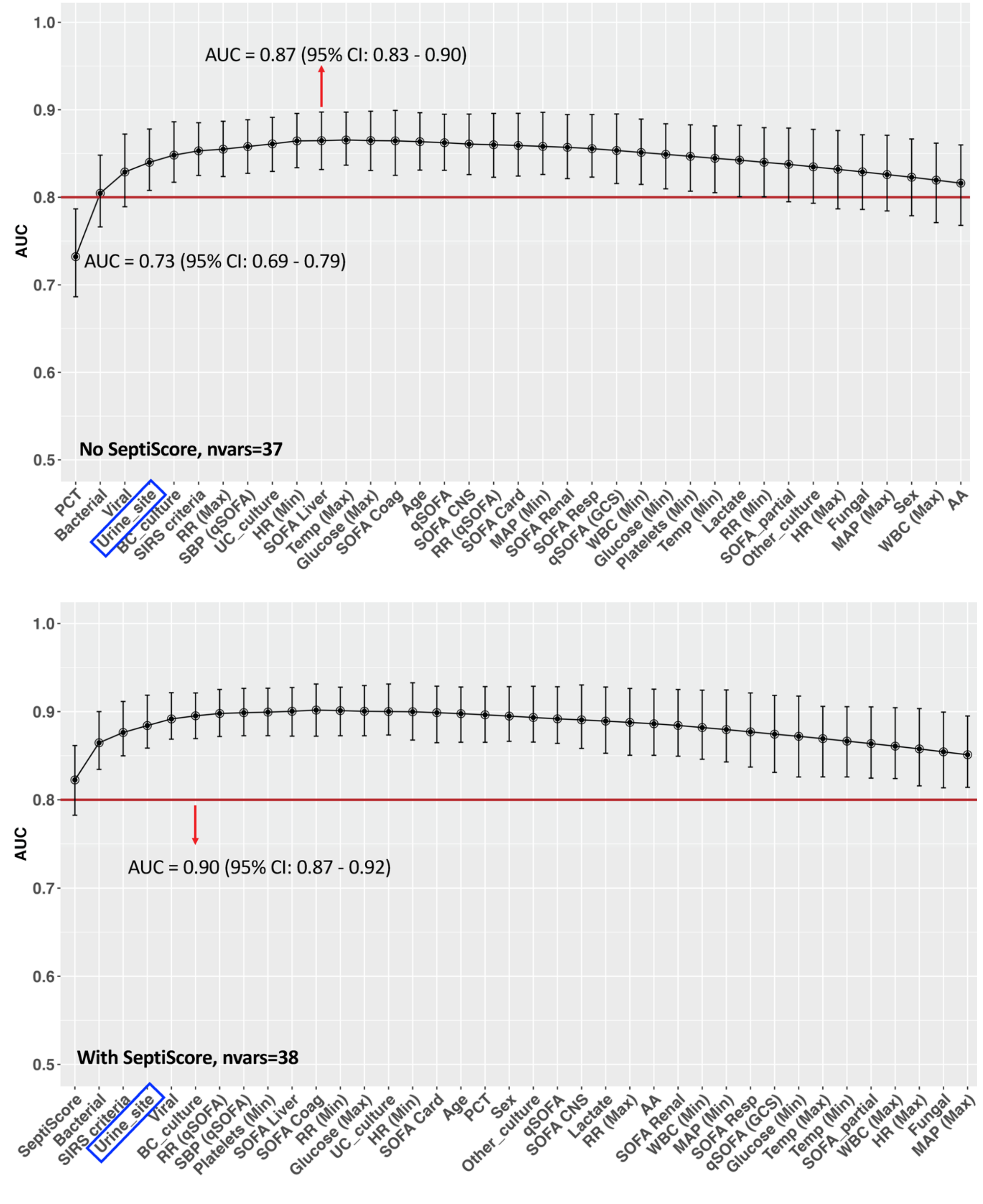
AUC
1.0
0.9
0.8
0.7
0.6
0.5
AUC = 0.87 (95% CI: 0.83 - 0.90)
AUC = 0.73 (95% CI: 0.69 - 0.79)
No SeptiScore, nvars=37
PCT
Bacterial
Viral
Urine_site
BC_culture
SIRS criteria
RR (Max)
SBP (qSOFA)
UC_culture
HR (Min)
SOFA Liver
Temp (Max)
Glucose (Max)
SOFA Coag
Age
qSOFA
SOFA CNS
RR (qSOFA)
SOFA Card
MAP (Min)
SOFA Renal
SOFA Resp
qSOFA (GCS)
WBC (Min)
Glucose (Min)
Platelets (Min)
Temp (Min)
Lactate
RR (Min)
SOFA_partial
Other_culture
HR (Max)
Fungal
MAP (Max)
Sex
WBC (Max)
AA
AUC
1.0
0.9
0.8
0.7
0.6
0.5
AUC = 0.90 (95% CI: 0.87 - 0.92)
With SeptiScore, nvars=38
SeptiScore
Bacterial
SIRS criteria
Urine_site
Viral
BC_culture
RR (qSOFA)
SBP (qSOFA)
Platelets (Min)
SOFA Liver
SOFA Coag
RR (Min)
Glucose (Max)
UC_culture
HR (Min)
SOFA Card
Age
PCT
Sex
Other_culture
qSOFA
SOFA CNS
Lactate
RR (Max)
AA
SOFA Renal
WBC (Min)
MAP (Min)
SOFA Resp
qSOFA (GCS)
Glucose (Min)
Temp (Max)
Temp (Min)
SOFA_partial
WBC (Max)
HR (Max)
Fungal
MAP (Max)

C: Blood (Bacteremia)

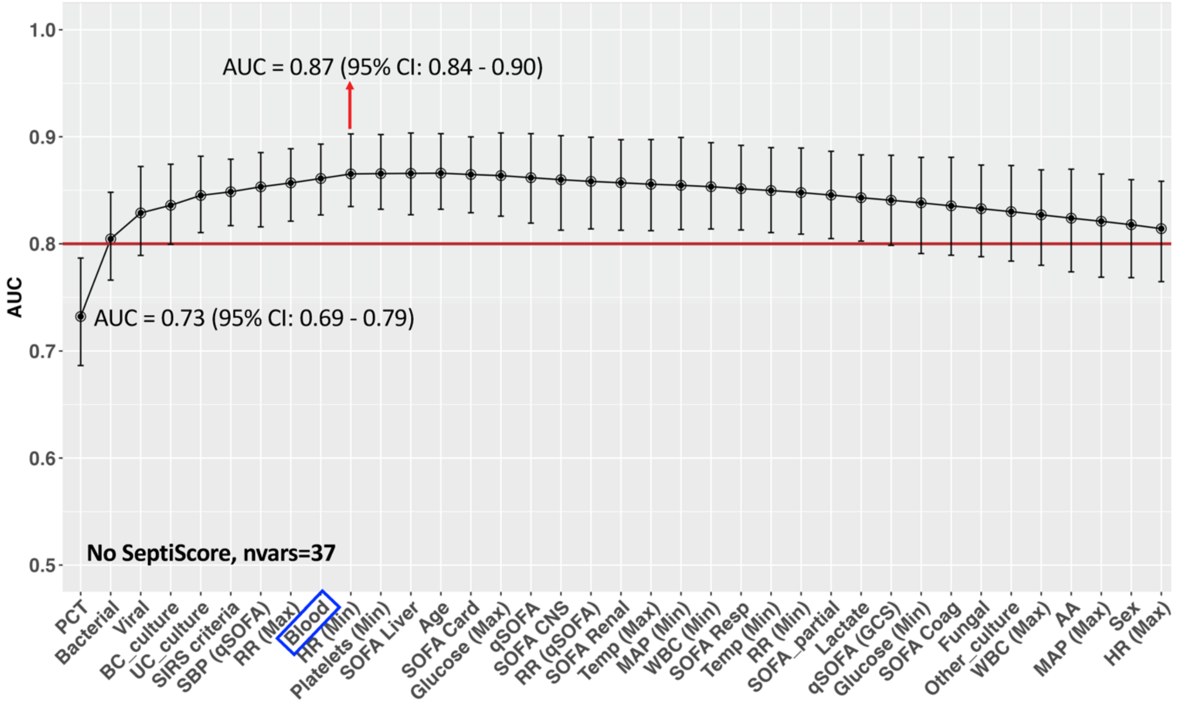


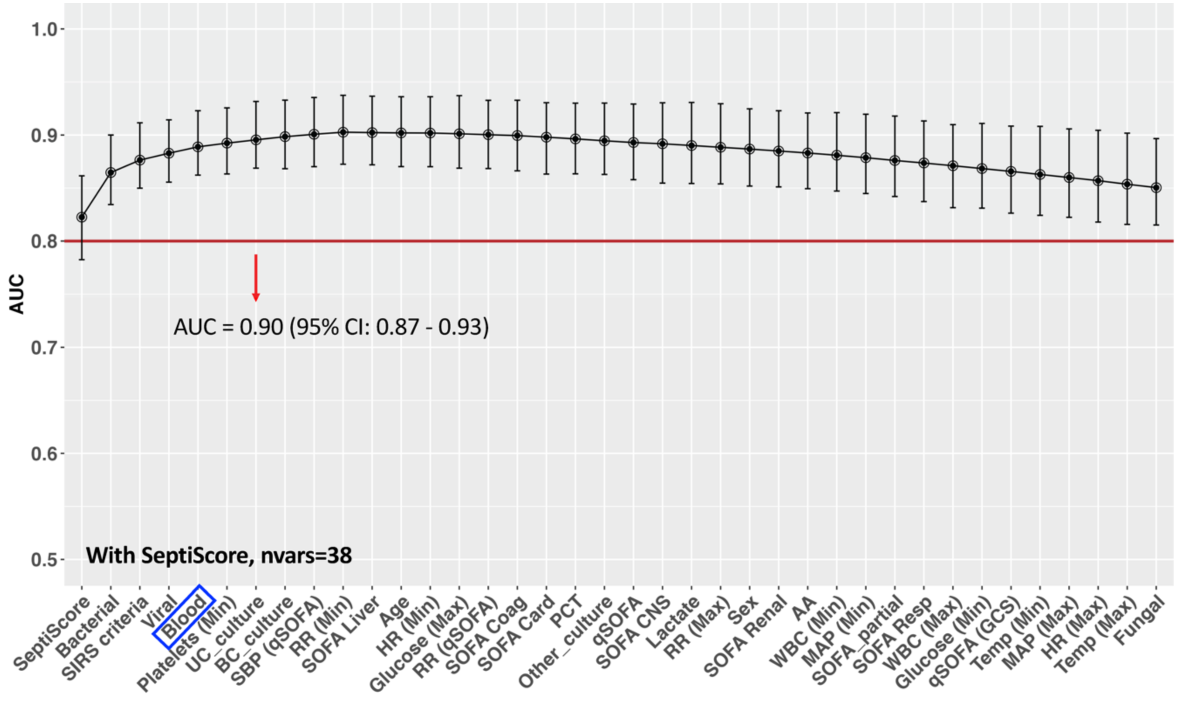

**D: CNS**

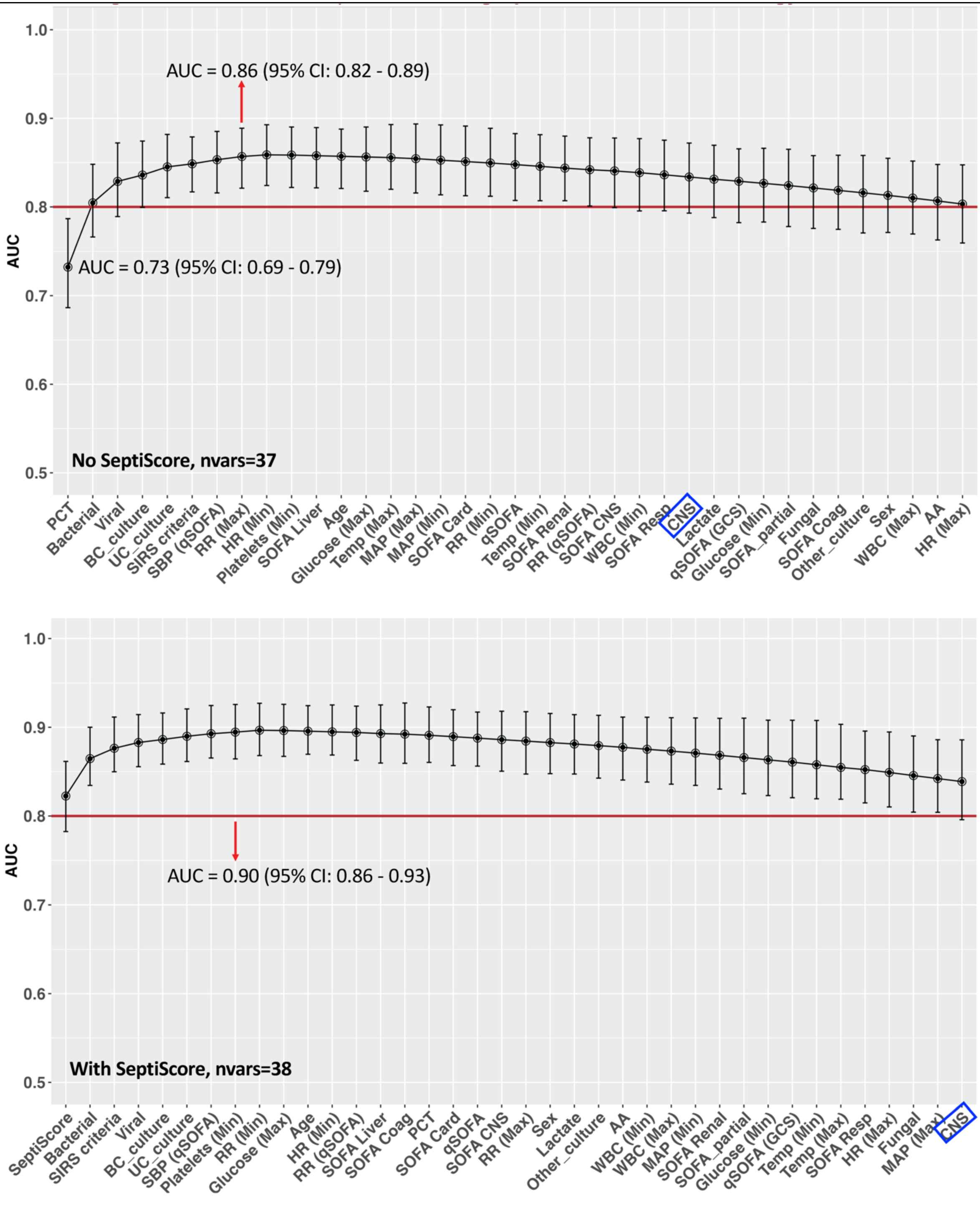
AUC = 0.86 (95% CI: 0.82 - 0.89)
AUC = 0.73 (95% CI: 0.69 - 0.79)
No SeptiScore, nvars=37
AUC
PCT
Bacterial
Viral
BC_culture
UC_culture
SIRS criteria
SBP (qSOFA)
RR (Max)
HR (Min)
Platelets (Min)
SOFA Liver
Age
Glucose (Max)
Temp (Max)
MAP (Max)
MAP (Min)
SOFA Card
RR (Min)
qSOFA
Temp (Min)
SOFA Renal
RR (qSOFA)
SOFA CNS
WBC (Min)
SOFA Resp
CNS
Lactate
qSOFA (GCS)
Glucose (Min)
SOFA_partial
Fungal
SOFA Coag
Other_culture
Sex
WBC (Max)
AA
HR (Max)
AUC = 0.90 (95% CI: 0.86 - 0.93)
With SeptiScore, nvars=38
AUC
SeptiScore
Bacterial
SIRS criteria
Viral
BC_culture
UC_culture
SBP (qSOFA)
Platelets (Min)
RR (Min)
Glucose (Max)
Age
HR (Min)
RR (qSOFA)
SOFA Liver
SOFA Coag
PCT
SOFA Card
qSOFA
SOFA CNS
RR (Max)
Sex
Lactate
Other_culture
AA
WBC (Min)
WBC (Max)
MAP (Min)
SOFA Renal
SOFA_partial
Glucose (Min)
qSOFA (GCS)
Temp (Min)
Temp (Max)
SOFA Resp
HR (Max)
Fungal
MAP (Max)
CNS

**E: Other Site of Infection**

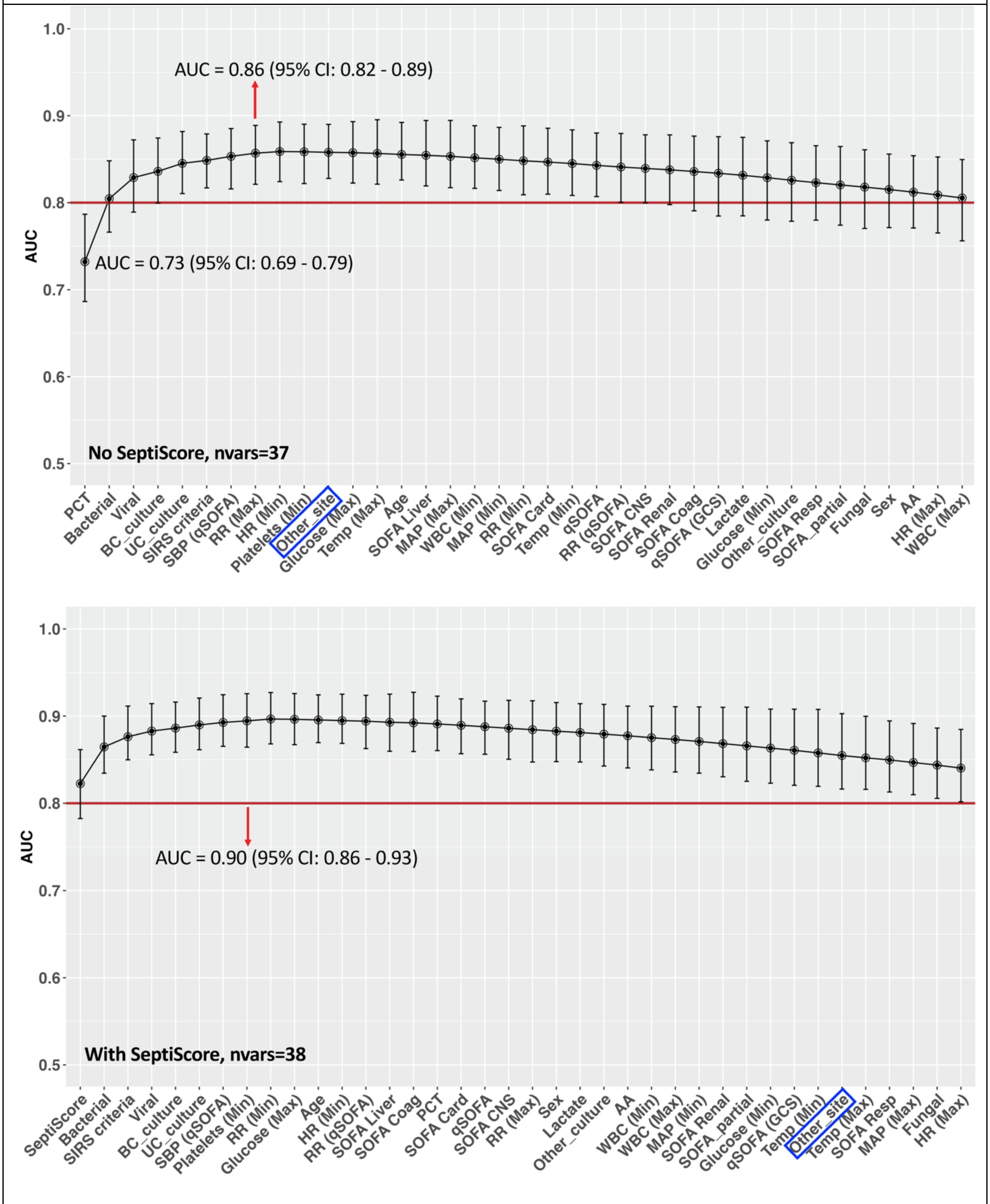

**F: No Site of Infection identified/ documented at Initial Impression**

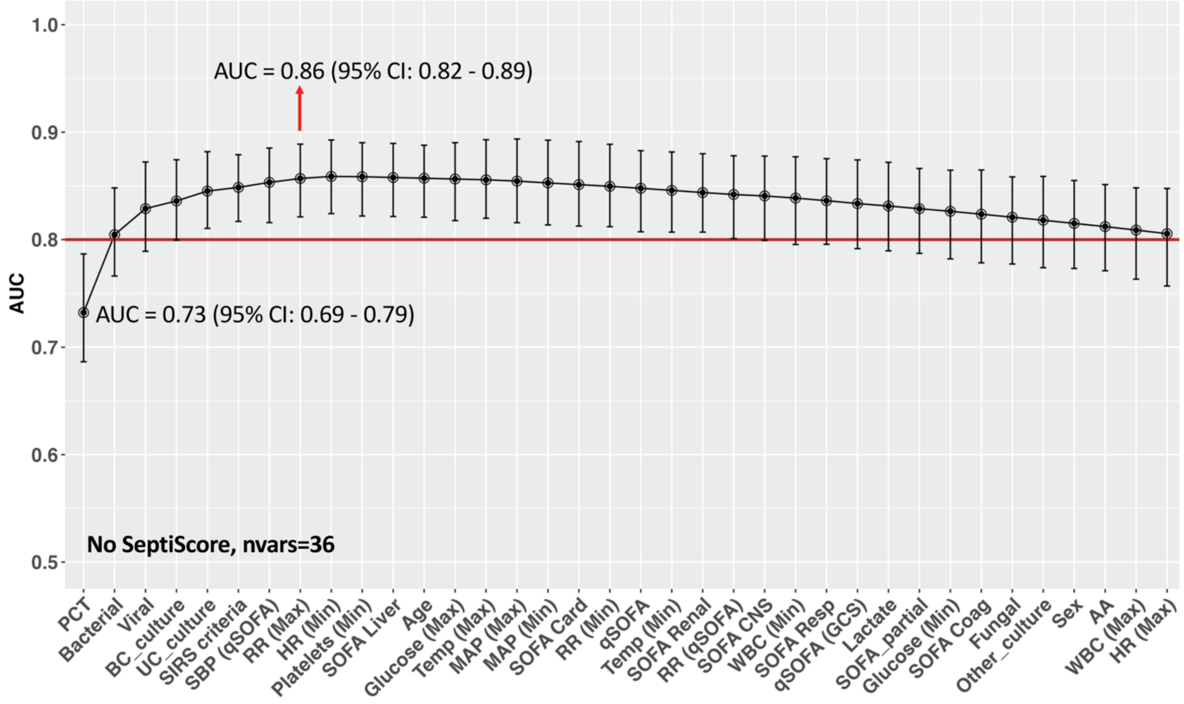


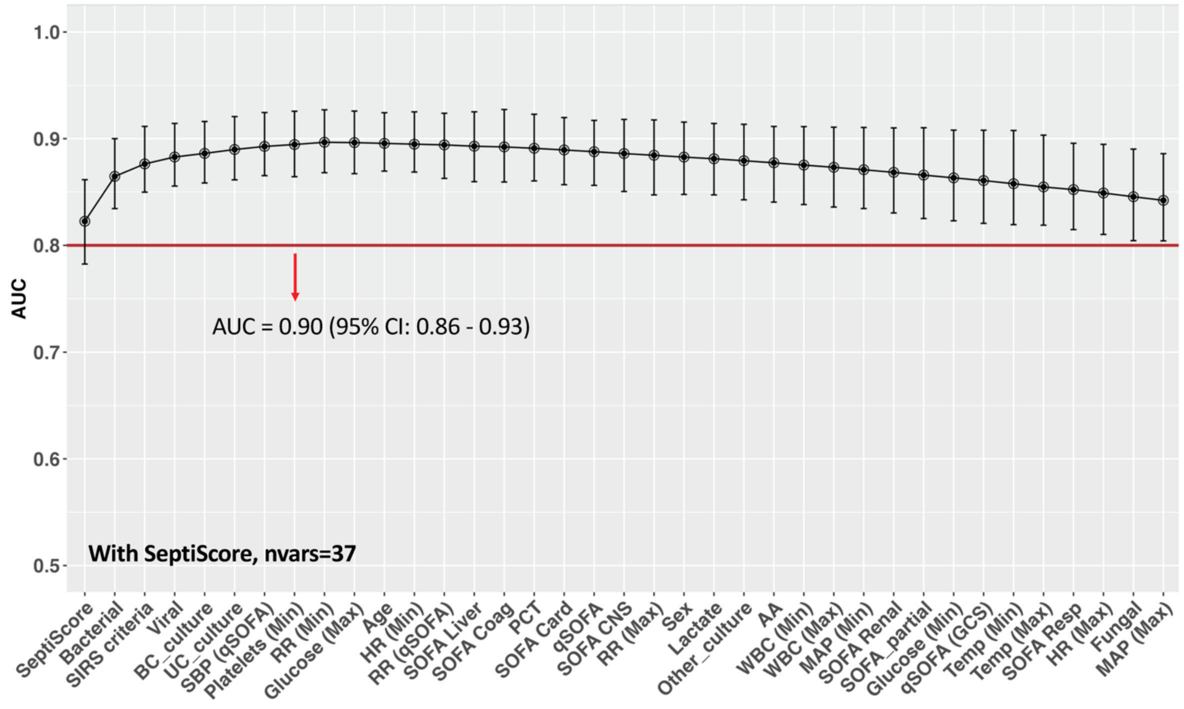

**Supplementary Table S4. Greedy Search Results for the Andalusian Cohort, at Time T1 (Presentation).**

| Source of infection | Starting Mean AUC (95% CI) | Maximum possible Mean AUC (95% CI) | Variable 1 | Variable 2 | Variable 3 | Variable 4 | Variable 5 | Variables sorted by order of importance | No. of variables needed to reach max performance |
|---|---|---|---|---|---|---|---|---|---|
| No site of infection | 0.648 (0.585 - 0.731) | 0.671 (0.616 - 0.738) | DBP | HR | Sex | | | DBP, HR, Sex | 3 |
| Pulmonary | | 0.671 (0.616 - 0.738) | DBP | HR | Sex | | | DBP, HR, Sex | 3 |
| Abdominal | | 0.671 (0.616 - 0.738) | DBP | HR | Sex | | | DBP, HR, Sex | 3 |
| Bacteremia | | 0.68 (0.63 - 0.741) | DBP | HR | Sex | Blood | | DBP, HR, Sex, Blood | 4 |
| Urine_site | | 0.671 (0.616 - 0.738) | DBP | HR | Sex | | | DBP, HR, Sex | 3 |
| CNS | | 0.671 (0.616 - 0.738) | DBP | HR | Sex | | | DBP, HR, Sex | 3 |
| Other site | | 0.68 (0.625 - 0.742) | DBP | HR | Other site | Sex | | DBP, HR, Other site, Sex | 4 |

**Supplementary Table S5. Greedy Search Results for the Andalusian Cohort at Time T2 (1-3 Hours Post-Presentation) Without SeptiCyte RAPID.**

| Source of infection | Starting Mean AUC (95% CI) | Maximum possible Mean AUC (95% CI) | Variable 1 | Variable 2 | Variable 3 | Variable 4 | Variable 5 | Variables sorted by order of importance | No. of variables needed to reach max performance |
|---|---|---|---|---|---|---|---|---|---|
| No site of infection | 0.779 (0.727 - 0.836) | 0.799 (0.75 - 0.848) | PCT | CRP | DBP | Creatinine | | PCT, CRP, DBP, Creatinine | 4 |
| Pulmonary | | 0.799 (0.75 - 0.848) | PCT | CRP | DBP | Creatinine | | PCT, CRP, DBP, Creatinine | 4 |
| Abdominal | | 0.799 (0.75 - 0.848) | PCT | CRP | DBP | Creatinine | | PCT, CRP, DBP, Creatinine | 4 |
| Bacteremia | | 0.799 (0.75 - 0.848) | PCT | CRP | DBP | Creatinine | | PCT, CRP, DBP, Creatinine | 4 |
| Urine | | 0.799 (0.75 - 0.848) | PCT | CRP | DBP | Creatinine | | PCT, CRP, DBP, Creatinine | 4 |
| CNS | | 0.799 (0.75 - 0.848) | PCT | CRP | DBP | Creatinine | | PCT, CRP, DBP, Creatinine | 4 |
| Other site | | 0.808 (0.757 - 0.859) | PCT | Other site | CRP | DBP | Creatinine | PCT, Other site, CRP, DBP, Creatinine, Platelets, Eosinophils, Sex, HR, RR | 10 |

**Supplementary Table S6. Greedy Search Results for the Andalusian Cohort at Time T2 (1-3 Hours Post-Presentation) With SeptiCyte RAPID.**

| Source of infection | Starting Mean AUC (95% CI) | Maximum possible Mean AUC (95% CI) | Variable 1 | Variable 2 | Variable 3 | Variable 4 | Variable 5 | Variables sorted by order of importance | No. of variables needed to reach max performance |
|---|---|---|---|---|---|---|---|---|---|
| No site of infection | 0.839 (0.786 - 0.902) | 0.867 (0.825 - 0.913) | SeptiScore | DBP | Age | Creatinine | | SeptiScore, DBP, Age, Creatinine | 4 |
| Pulmonary | | 0.867 (0.825 - 0.913) | SeptiScore | DBP | Age | Creatinine | | SeptiScore, DBP, Age, Creatinine | 4 |
| Abdominal | | 0.867 (0.825 - 0.913) | SeptiScore | DBP | Age | Creatinine | | SeptiScore, DBP, Age, Creatinine | 4 |
| Bacteremia | | 0.867 (0.825 - 0.913) | SeptiScore | DBP | Age | Creatinine | | SeptiScore, DBP, Age, Creatinine | 4 |
| Urine | | 0.867 (0.825 - 0.913) | SeptiScore | DBP | Age | Creatinine | | SeptiScore, DBP, Age, Creatinine | 4 |
| CNS | | 0.867 (0.825 - 0.913) | SeptiScore | DBP | Age | Creatinine | | SeptiScore, DBP, Age, Creatinine | 4 |
| Other site | | 0.867 (0.825 - 0.913) | SeptiScore | DBP | Age | Creatinine | | SeptiScore, DBP, Age, Creatinine | 4 |

**Supplementary Table S7. Greedy Search Results for the Andalusian Cohort at Time T3 (1-3 Days Post-Presentation) Without SeptiCyte RAPID.**

| Source of infection | Starting Mean AUC (95% CI) | Maximum possible Mean AUC (95% CI) | Variable 1 | Variable 2 | Variable 3 | Variable 4 | Variable 5 | Variables sorted by order of importance | No. of variables needed to reach max performance |
|---|---|---|---|---|---|---|---|---|---|
| No site of infection | 0.784 (0.76 - 0.812) | 0.895 (0.864 - 0.924) | Bacterial infection | CRP | DBP | Fungal infection | | Bacterial infection, CRP, DBP, Fungal infection | 4 |
| Pulmonary | | 0.895 (0.864 - 0.924) | Bacterial infection | CRP | DBP | Fungal infection | | Bacterial infection, CRP, DBP, Fungal infection | 4 |
| Abdominal | | 0.895 (0.864 - 0.924) | Bacterial infection | CRP | DBP | Fungal infection | | Bacterial infection, CRP, DBP, Fungal infection | 4 |
| Bacteremia | | 0.895 (0.864 - 0.924) | Bacterial infection | CRP | DBP | Fungal infection | | Bacterial infection, CRP, DBP, Fungal infection | 4 |
| Urine | | 0.895 (0.864 - 0.924) | Bacterial infection | CRP | DBP | Fungal infection | | Bacterial infection, CRP, DBP, Fungal infection | 4 |

| Source of infection | Starting Mean AUC (95% CI) | Maximum possible Mean AUC (95% CI) | Variable 1 | Variable 2 | Variable 3 | Variable 4 | Variable 5 | Variables sorted by order of importance | No. of variables needed to reach max performance |
|---|---|---|---|---|---|---|---|---|---|
| CNS | | 0.895 (0.864 - 0.924) | Bacterial infection | CRP | DBP | Fungal infection | | Bacterial infection, CRP, DBP, Fungal infection | 4 |
| Other site | | 0.906 (0.877 - 0.931) | Bacterial infection | CRP | DBP | Fungal infection | Other site | Bacterial infection, CRP, DBP, Fungal infection, Other site, BC Positive, Creatinine | 7 |

**Supplementary Table S8. Greedy Search Results for the Andalusian Cohort at Time T3 (1-3 Days Post-Presentation) With SeptiCyte RAPID.**

| Source of infection | Starting Mean AUC (95% CI) | Maximum possible Mean AUC (95% CI) | Variable 1 | Variable 2 | Variable 3 | Variable 4 | Variable 5 | Variables sorted by order of importance | No. of variables needed to reach max performance |
|---|---|---|---|---|---|---|---|---|---|
| No site of infection | 0.839 (0.786 - 0.902) | 0.927 (0.9 - 0.949) | SeptiScore | Bacterial infection | DBP | Blood culture positive | | SeptiScore, Bacterial infection, DBP, Blood Culture Positive | 4 |
| Pulmonary | | 0.927 (0.9 - 0.949) | SeptiScore | Bacterial infection | DBP | Blood culture positive | | SeptiScore, Bacterial infection, DBP, Blood Culture Positive | 4 |
| Abdominal | | 0.927 (0.9 - 0.949) | SeptiScore | Bacterial infection | DBP | Blood culture positive | | SeptiScore, Bacterial infection, DBP, Blood Culture Positive | 4 |
| Bacteremia | | 0.927 (0.9 - 0.949) | SeptiScore | Bacterial infection | DBP | Blood culture positive | | SeptiScore, Bacterial infection, DBP, Blood Culture Positive | 4 |

| Source of infection | Starting Mean AUC (95% CI) | Maximum possible Mean AUC (95% CI) | Variable 1 | Variable 2 | Variable 3 | Variable 4 | Variable 5 | Variables sorted by order of importance | No. of variables needed to reach max performance |
|---|---|---|---|---|---|---|---|---|---|
| Urine | | 0.927 (0.9 - 0.949) | SeptiScore | Bacterial infection | DBP | Blood culture positive | | SeptiScore, Bacterial infection, DBP, Blood Culture Positive | 4 |
| CNS | | 0.927 (0.9 - 0.949) | SeptiScore | Bacterial infection | DBP | Blood culture positive | | SeptiScore, Bacterial infection, DBP, Blood Culture Positive | 4 |
| Other site | | 0.936 (0.911 - 0.956) | SeptiScore | Bacterial infection | DBP | Other site | Fungal infection | SeptiScore, Bacterial infection, DBP, Other site, Fungal infection | 5 |

**Supplementary Figure S4 (A-B).** Investigation of potential incorporation bias, by comparative ROC curve analysis. **(A)** Baseline ROC curve for seven variables (AUC 0.85, 95% CI 0.80-0.89), and ROC curves generated by removing variables individually. The seven variables were SeptiCyte RAPID, PCT, Temperature (max), RR (max), MAP (max), age, and HR (min). The orange curve (AUC 0.75, 95% CI 0.70-0.79) results from removing SeptiScore from the logistic equation. Removal of other variables individually have only small detrimental effects. **(B)** ROC curve for SeptiScore + PCT (not subject to incorporation bias), with AUC 0.83 (95% CI 0.78-0.87; purple curve). Removal of PCT reduces the AUC to 0.82 (95% CI 0.78-0.86; green curve), while removal of SeptiScore reduces the AUC to 0.73 (95% CI 0.68-0.78; orange curve).

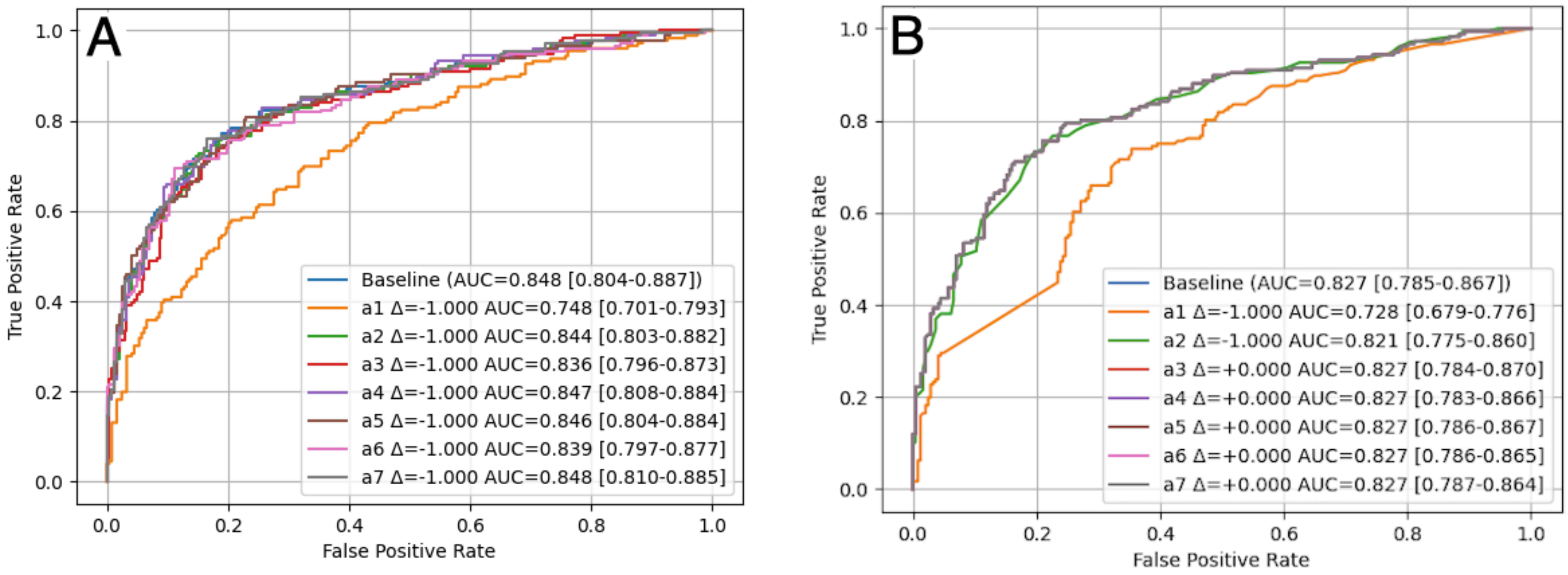

**Supplementary Table S9.** Relative magnitude of contributions of logistic regression input variables, arranged in decreasing order of impact upon the output AUC. Change in AUC is measured relative to the baseline logistic regression fit using all seven variables (AUC 0.85; 95% CI 0.80-0.89).

| Variable | Subject to Incorporation Bias? | Logistic Regression Coefficient | AUC when coefficient(s) set to zero | AUC 95% CI | Change in AUC |
|---|---|---|---|---|---|
| x1 SeptiScore | No | 0.6375 | 0.75 | 0.70-0.89 | -0.10 |
| x2 PCT | | 0.0154 | 0.84 | 0.80-0.88 | -0.01 |
| x1 & x2 (jointly) | | | 0.71 | 0.66-0.76 | -0.14 |
| x3 Temp (max) | Yes | 0.6268 | 0.84 | 0.80-0.87 | -0.01 |
| x4 RR (max) | | 0.0366 | 0.85 | 0.81-0.88 | -0.00 |
| x5 MAP (max) | | -0.0091 | 0.85 | 0.80-0.88 | -0.00 |
| x6 Age | | 0.0191 | 0.84 | 0.80-0.88 | -0.01 |
| x7 HR (min) | | 0.0073 | 0.85 | 0.81-0.88 | -0.00 |

Of the variable subject to incorporation bias, exclusion of Temperature (max) produced the largest reduction in AUC (Δ AUC = −0.01), while exclusion of the remaining incorporation bias-sensitive variables resulted in smaller changes (Δ AUC < −0.01). These Δ AUC values represent upper bounds on the combined effect of each variable's legitimate predictive contribution and any inflation due to incorporation bias. Importantly, no single adjudication-incorporated variable (x3-x7) accounted for more than approximately 0.01 AUC units of the model's discrimination of sepsis vs. SIRS. This indicates that the observed AUC is not strongly driven by any individual incorporated variable and is relatively robust to their exclusion. While this analysis does not eliminate or quantify incorporation bias in absolute terms, it suggests that incorporation-related inflation is unlikely to be dominated by any single variable.

We additionally examined the joint effect of excluding all adjudication-incorporated variables by retaining only x1 (SeptiScore) and x2 (PCT) in the model. When the coefficients for x3–x7 were simultaneously set to zero, the resulting model yielded an AUC of 0.83 (95% CI 0.78-0.87), representing a reduction of 0.02 relative to the full seven-variable model.

This joint-exclusion analysis provides an upper bound on the combined contribution of variables subject to incorporation bias, encompassing both their legitimate predictive information and any inflation arising from their use in adjudication. Notably, the majority of the model's discriminative performance was preserved using only variables independent of the reference diagnosis, indicating that apparent discrimination is not primarily driven by adjudication-incorporated inputs.
